# Disease control as an optimization problem

**DOI:** 10.1101/2020.09.15.20194811

**Authors:** Miguel Navascués, Costantino Budroni, Yelena Guryanova

## Abstract

Traditionally, expert epidemiologists devise policies for disease control through a mixture of intuition and brute force. Namely, they use their know-how to narrow down the set of logically conceivable policies to a small family described by a few parameters, following which they conduct a grid search to identify the optimal policy within the set. This scheme is not scalable, in the sense that, when used to optimize over policies which depend on many parameters, it will likely fail to output an optimal disease policy in time for its implementation. In this article, we use techniques from convex optimization theory and machine learning to conduct optimizations over disease policies described by hundreds of parameters. In contrast to past approaches for policy optimization based on control theory, our framework can deal with arbitrary uncertainties on the initial conditions and model parameters controlling the spread of the disease. In addition, our methods allow for optimization over weekly-constant policies, specified by either continuous or discrete government measures (e.g.: lockdown on/off). We illustrate our approach by minimizing the total time required to eradicate COVID-19 within the Susceptible-Exposed-Infected-Recovered (SEIR) model proposed by Kissler *et al*. (March, 2020).

## I. INTRODUCTION

At the time of writing, the COVID-19 pandemic had already caused more than half a million deaths worldwide; at the time of re-writing, some weeks later, this number was approaching one million. The effects of the virus have been widespread and substantial, from the collapse of healthcare systems [1-3] to the enforcement of isolation and quarantine. In the case of Nepal, the national lockdown lasted for 120 days uninterrupted [4].

In these circumstances, identifying reliable and effective disease control policies is of utmost importance. Here by “policy” we mean a deliberate intervention intended to mitigate the effects of a disease as it runs its course. In much of the mathematical literature on epidemiology, the process of generating a policy is as follows [5]: (1) based on their intuition, expert epidemiologists propose a number of suitable policies to control the disease; (2) the impact on the population of each of the considered policies is assessed through dynamical models of disease spread; (3) the outcomes of all policies are compared and a decision is taken as to which one is deemed to be the best.

This three-step process has two disadvantages. First of all, the class of policies devised by an expert could well be suboptimal, since the optimal policy (under some figure of merit) could be extremely complicated and counterintuitive. Second, the method requires one to numerically simulate each policy. Given the exponential growth of the number of policies in the number of control parameters, the number of policies considered may be on the order of billions; hence, this strategy is not guaranteed to identify the optimal disease control policy in time to enforce it.

More sophisticated approaches for disease control rely on optimal control theory to identify a suitable policy (see, e.g., [6-9]). The starting point of all these works is that both the initial conditions (namely, the number of infected, exposed, etc.) and the model parameters (such as the disease’s reproduction number) specifying the spread of the disease are known with high precision. This requirement is never met in a real-life epidemic, especially close to the outbreak, when the uncertainty of the disease’s reproduction number can be very high [10, 11]. Policies derived through optimal control theory are thus not guaranteed to have the desired effects in practice.

An additional disadvantage of optimal control theory is that government measures in the model cannot be constrained to be discrete, such as a lockdown which is either on or off, a policy that so far has dominated global efforts to control the COVID-19 epidemic [12]. Last but not least, optimal control theory can only handle scenarios where the policies continuously vary over time, thus not making it compatible with observed government measures to control COVID-19, which for the most part have been applied on a weekly basis.

In this paper we introduce a general framework that maps any disease control scenario to an optimization problem. Contrary to the optimal control approach, our framework can accommodate constraints on the disease dynamics which must hold for whole regions of the initial conditions and the disease’s model parameters. Our framework can enforce policies to be weekly and/or discrete. Invoking tools from optimization theory and machine learning [13], we propose efficient heuristics to solve the optimization problem and hence identify the government policy that best controls the disease.

To illustrate the power of our approach, we use these optimization techniques to generate long-term plans to fight COVID-19, under the assumption that the disease’s dynamics are accurately captured by a variant of the Susceptible-Exposed-Infected-Recovered (SEIR) compartmental model [5] proposed in [14]. Our results confirm that optimal policies tend to be too complicated to be devised by a human.

In reality, most epidemiological models only provide short-term approximations to the spread of the disease, with long term projections becoming less and less reliable [15]. In addition, notwithstanding the enormous knowledge gathered since the initial COVID-19 outbreak, many questions remain to be answered regarding the correctness and accuracy of compartmental models such as SEIR: their basic assumptions (e.g., are recovered patients temporarily or permanently immune to the disease?); the actual value of the model’s parameters (e.g., the basic reproduction number *R*_0_); and the role of variables not modeled (e.g., age, geographic distribution, contact tracing policies, role of superspreaders).

In this regard, the goal of this work is not to propose a concrete government policy, but rather to present an efficient method to obtain an optimal one, given all the available information. To estimate the effect of our methods in a realistic scenario, we conduct a numerical simulation where we re-calculate the optimal policy plans every month, based on new, incoming data. The very final policy plan that we present requires less stringent physical distancing measures compared with the one which is not re-calculated every month. All the code used in our simulations is freely available at[16].

## II. THE FRAMEWORK

Our starting point is an epidemic that affects a closed population. This assumption is not limiting, since a large ensemble of population centres where individuals are free to commute can also be modeled as a closed system [17]. To gain an understanding of how the disease spreads, it is standard to divide the population into different sectors or compartments [5] (see Fig. 1). One can define, e.g., the compartment of all those individuals who are currently infected. This compartment can, in turn, be sub-divided into different compartments, such as symptomatic/asymptomatic. Once the number of relevant compartments is fixed, one can estimate the occupation of each of them and arrange the resulting numbers in a vector ***x***. The disease is subsequently analysed by looking at how ***x*** changes with time.

**FIG. 1:**
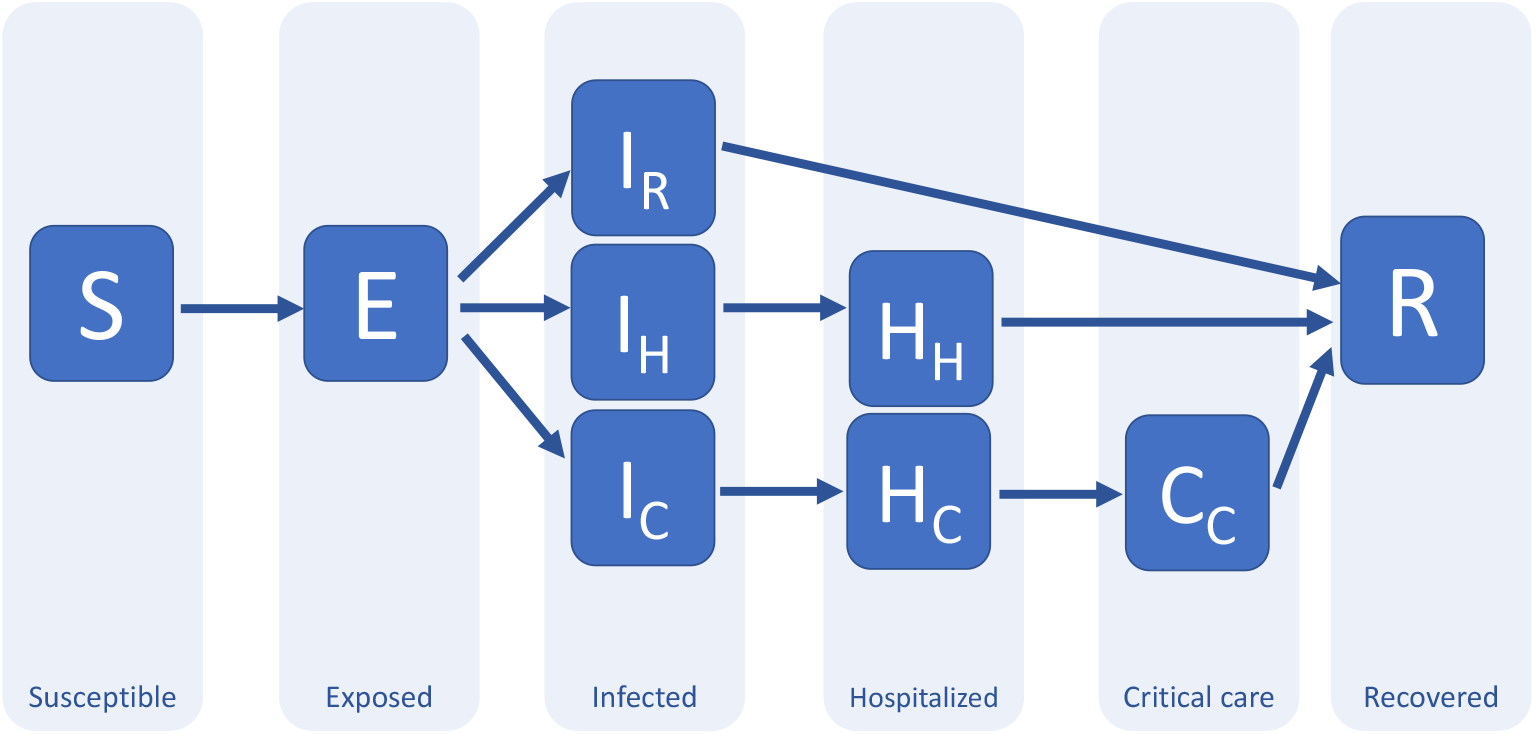
A possible compartment model for COVID-19 (adapted from [14]). The main compartments are: susceptible; exposed; infected; hospitalized; critical and recovered. This compartmental splitting captures different possible evolutions as well as time delays between transitions. The “infected” compartment, for instance, contains those who will recover without hospitalization (*I*_*R*_); those who will be hospitalized but won’t need critical care (*I*_*H*_); and those who will end up receiving critical care (*I*_*C*_). The “exposed” compartment is introduced here to model the time delay between the exposure to the disease and the development of symptoms (incubation period), in particular, the possibility of infecting others, which is what is relevant for the model. In this model, the compartment “recovered” includes both dead and alive individuals; in principle, it could be sub-divided further.

In order to control or even extinguish an epidemic, governments can enforce a number of different measures: mass vaccination, physical (or social [18]) distancing measures, or even a full lockdown, are common examples of interventions aimed at fighting the disease. When and to which degree such measures are applied is determined by the *disease control policy*. Consider a disease control policy based on random vaccination campaigns, where the intervention consists of vaccinating a number of individuals at random per day. Call *v*(*t*) the number of individuals vaccinated on day *t*. This function, between the initial and final times *t*_0_ and *t*_*f*_ (i.e. on the interval [*t*_0_, *t*_*f*_]) determines the government’s vaccination policy. Similarly, let *s*(*t*) take the value 1 if the country is in lockdown on day *t* and 0 if it is not. Then the government’s lockdown policy between times *t*_0_ and *t*_*f*_ corresponds to the function *s*(*t*).

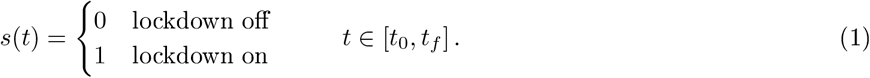

If the government is intervening both through vaccination and lockdown, then its disease control policy will be identified by *both* the functions *v*(*t*) and *s*(*t*).

The above are instances of *non-adaptive policies* for disease control, because the functions *s, v* just depend on the time *t*, and not, e.g., on the current value of the death toll. A general (adaptive) policy for disease control would take into account the whole past history of data gathered by the government before deciding what to do at each step. Although in the following all our proposed policies are non-adaptive, the formalism we introduce allows one to optimize over adaptive policies as well.

In conclusion, a disease control policy can always be identified with a vector function ***α*** of the time *t* and perhaps some other observed variables ***o***, where each vector entry represents a type of government intervention at time *t*. In turn, we can use a variable vector ***µ*** *∈ ℝ*^*n*^to parametrize the class of considered policies, that is, ***α***(*t*, ***o***) = ***α***(*t*, ***o***; ***µ***). Since ***µ*** completely determines the policy ***α***, we can also regard the parameters ***µ*** as the disease policy. We will do so from now on.

The applied policy ***µ*** is assumed to influence the compartment occupation within the time interval [*t*_0_, *t*_*f*_]. That is, ***x*** is both a function of *t* and ***µ***. In this work we are interested in devising policies for disease control which guarantee that the spread of the disease evolves under certain conditions. For example, any country has a fixed number of critical care capacity beds, which we denote by *B*_*c*_. At each time *t ∈* [*t*_0_, *t*_*f*_], it is desirable that the number of individuals admitted to critical care in hospitals, *𝒞* (*t*), does not exceed that capacity. That is, we require that

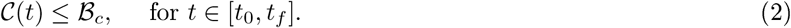

We will call any such condition on the evolution of the *disease a constraint*.

Finally, among all policies satisfying the desired constraints, we typically wish to identify the one that minimizes a certain quantity. For example, for many diseases, a simple physical distancing policy that satisfies the constraint (2) consists of declaring a lockdown throughout the whole time interval [*t*_0_, *t*_*f*_], i.e., *s*(*t*) = 1 for *t ∈* [*t*_0_, *t*_*f*_]. This policy is arguably impractical, difficult to enforce and harmful to its citizens’ psychological health as well as to the national economy. More rationally, one is interested in finding alternative disease policies which, while respecting the critical care occupation constraint, minimize the number of days of lockdown. Alternatively, one may wish to minimize the total number of deaths during the interval [*t*_0_, *t*_*f*_], or the number of infected people. In general, the figure of merit, or quantity that we wish to minimize will be a complicated functional *f* of the considered policy ***α*** and ***x***. We will call this functional the *objective function*.

## III. MODELS AND OPTIMIZATION

The optimization problem sketched above is mathematically ill-defined, unless we specify how ***x*** varies with the parameters ***µ*** determining the policy. In order to predict the natural course of a disease or how a given policy might affect its spread, epidemiologists make use of mathematical models. In this paper, we will mainly be concerned with *deterministic compartmental models*. In these models, the whole population is divided into a number of basic compartments and the interactions between those compartments are modeled through a system of ordinary differential equations. Given the occupation ***x***_**0**_ of the compartments at time *t*_0_, these models allow us to compute the value of ***x*** at any instant *t ∈* [*t*_0_, *t*_*f*_] as a function of the policy ***µ***. That is, each model provides an implicit functional relation of the form ***x*** = ***x***(*t*; ***µ, x***_**0**_).

Past literature on disease control has made extensive use of compartmental models to recommend specific strategies for policy-makers in an effort to control the spread of disease. In many cases, suitable policies are devised through a mixture of intuition and grid search, see [14, 19-21]. The starting point is a family of disease control policies with one or two unknowns. For instance, in pulse vaccination [20], those unknowns are the time intervals between two random mass vaccinations and the vaccination rate. In this paper, we propose a scheme that allows for the optimization over policies specified by thousands of parameters in the space of a few hours. Our scheme, detailed in Appendices B and C, is based on a standard tool in optimization theory and machine learning known as *gradient descent* [22, 23]. Starting with a rough guess for the optimal policy ***µ***^(0)^, gradient descent methods generate a sequence of policies ***µ***^(1)^, ***µ***^(2)^, … which typically exhibit increasingly better performance. Although the gradient method is not guaranteed to converge to the optimal policy, after many iterations it generates solutions that are good enough for many practical problems. In fact, gradient descent is the method most commonly used to train deep neural networks [13] and support vector machines [24].

Importantly, the computational resources required to carry out the gradient descent method are comparable to the cost of running a full simulation between times *t*_0_ and *t*_*f*_ with the considered disease model. Furthermore, the necessary computations can be *parallelized* for policies depending on many parameters. Although the focus of this paper is on compartmental disease models, our main ideas can also be used to understand ecological systems undergoing a more complex dynamics [25], see Appendix E. Even in such complicated scenarios, the former scaling laws hold: provided that we can run the considered disease model, we can apply the gradient method to optimize over policies of disease control.

To illustrate our approach for policy design, we will use a variant of the extended “susceptible-exposed-infected-recovered” (SEIR) disease model proposed in [14] to predict the impact of COVID-19 in the USA, a schematic of which is shown in Fig. (1) and the details of which appear in Appendix A. The clinical parameters of the model, such as recovery and hospitalization rates, were estimated in [26, 27], based on early reports from COVID-19 cases in the UK, China and Italy. Following the model in [14], the disease transmission is assumed to be seasonal by analogy with the known behavior of betacoronaviruses such as HCoV-OC 3 [27], with a baseline reproduction number between 2 and 2.5, following fits of the early growth-rate of the epidemic in Wuhan [10, 11].

A relevant compartment in this model is *𝒞* (*t*), the population occupying a critical care bed^1^ at time *t*. The patients sent to critical care cannot breathe unassisted, and thus it is fundamental to ensure that such capacity is not surpassed, namely, that the constraint (2) holds. For our simulations, we chose a population size of *N*_*pop*_ = 47 million and *B*_*c*_ = 9.5 *× N*_*pop*_ *×* 10^−5^. That is, we assumed that the healthcare system provides 95 critical care beds per million inhabitants. This is a good approximation to the healthcare capacity of many European countries, as well as the USA.

According to the chosen model, without intervention the number of citizens requiring a bed in a critical care unit evolves according to Figure 2 (see Appendix A for the exact initial conditions of our numerical simulations). As the reader can appreciate, between the third and seventh month, the number of people in need of critical care exceeds the capacity of the considered healthcare system by 18 times.

**FIG. 2:**
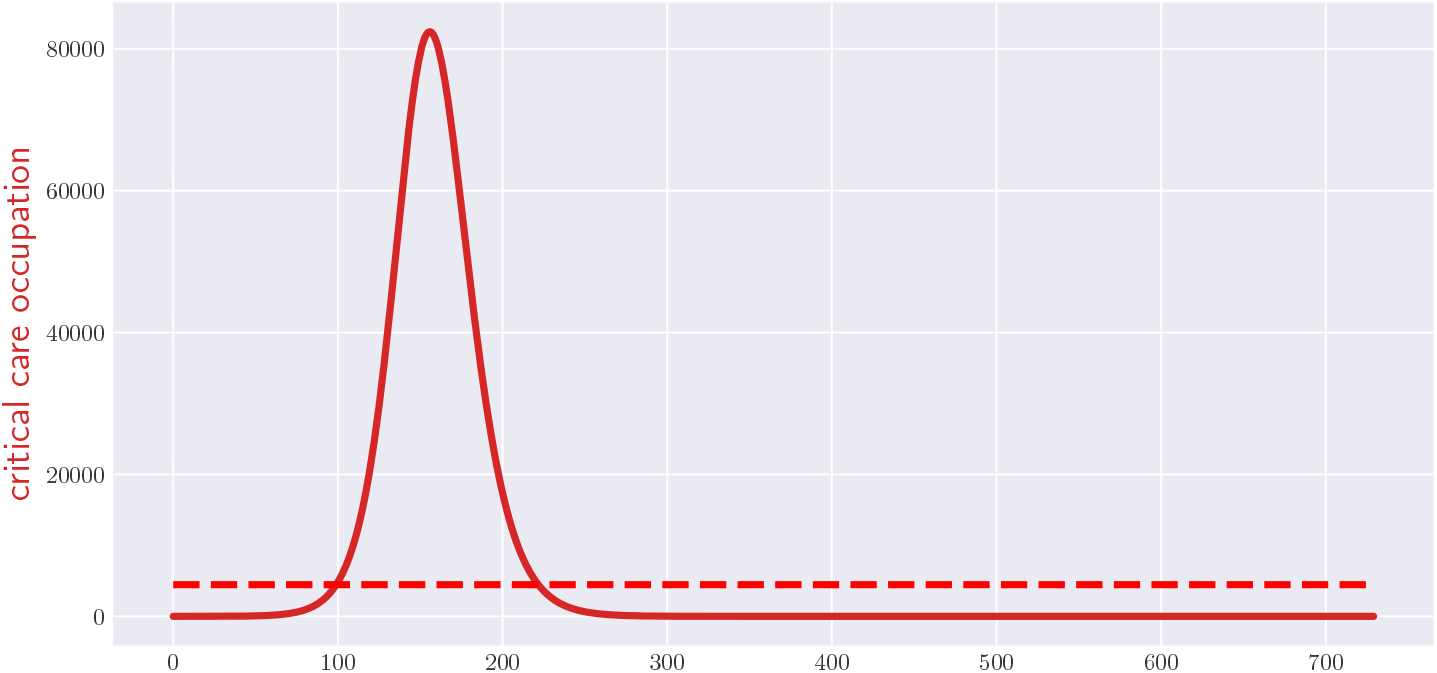
Occupation of critical care beds over two years with no policy intervention. The red dashed line indicates the critical care capacity of the healthcare system.

At the time of this writing, none of the potential vaccines for COVID-19 have passed the necessary clinical trials to be considered safe to administer [28]. In lieu of this, most governments have opted to control the disease via distancing measures and/or lockdowns. The effect of implementing a policy *s*(*t*) is to multiply the disease’s basic reproduction number *R*_0_ by a factor of *r* [14, 27], *i*.*e. R*_0_ *→ rR*_0_, where

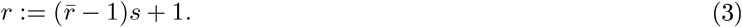

At this point we shall make a distinction: a policy *s* which outputs a binary value {0, 1} shall be known as *discrete* and correspond to situations in which a *lockdown* is either on or off. For discrete *s*(*t*) as in Eq. (1), the effect of a lockdown (*s* = 1) results in the reduction of the transmission rate 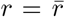 in Eq. (3). On the other hand, when the population is free to interact (*s* = 0) then *r* = 1 and there is no change in the disease’s basic reproduction number.

On the other hand, a policy *s*, which outputs a value on the interval [0, 1] will be known as *continuous* and correspond to *physical-distancing measures*. Continuous values of *s*(*t*) correspond to intermediate policies (for example mandatory face masks, suspension of sport events, remote working, school closures), the effect of which can be tentatively estimated from available data [26].

If distancing measures are the only type of intervention that a government uses, then a non-adaptive continuous policy is fully determined by the function *s*(*t*; ***µ***) *∈* [0, 1]. To begin with, we will assume that the government can only declare new measures at the beginning of each week, i.e., that *s*(*t*) does not vary within the weekly intervals *t ∈* [7*k*, 7(*k* + 1)] =: *I*_*k*_, for *k ∈* ℤ. With these conditions, *s*(*t*) can be expressed as

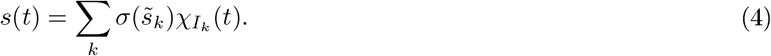

where *χI*_*k*_ (*t*) is the characteristic function of week number *k*, i.e., *χI*_*k*_ (*t*) equals 1 if *t* is in the *k*-th week; and 0, otherwise. The characteristic function, thus, ensures that there is only one policy *s*(*t*) per week. 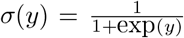 denotes the sigmoid function, which guarantees that *s*(*t*) ∊ [0, 1] by continuously mapping the variables 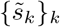, such that it is everywhere differentiable, making it amenable to the gradient method. The parameters to optimize over are 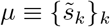, since they fully define the government’s disease policy.

Having chosen either discrete or continuous measures, one must choose a figure of merit or objective function to optimize over. Our formalism, explained in detail in Appendix C, allows us to minimize expressions of the form

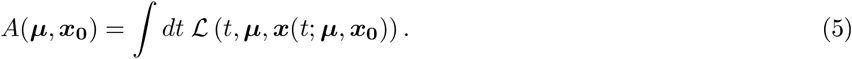

In this paper, we consider optimizations that never allow the number of people in the critical care compartment to exceed the maximal occupancy, i.e., we impose the constraint in Eq. (2). Thus we consider optimizations of the form

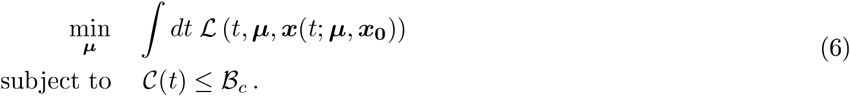

In what follows, we split our analysis by choosing different objective functions ℒ (*t*, ***µ, x***(*t*; ***µ, x***_**0**_)) to minimize over, considering policies *s*(*t*; ***µ***) which are either continuous, as in Eq. (4), or discrete as in Eq. (1).

### A. Objective function 1: minimizing physical distancing and lockdown measures

Suppose, for instance, that one is confident that a vaccine for COVID-19 will be developed within the next two years. In this case, one is interested in minimizing the aggregate economic cost *ε* associated with the physical-distancing measures implemented by the government over these two years. Thus for the optimization in Eq. (6), we consider Lagrangians (objective functions) of the form

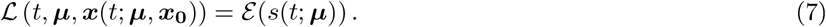

Figure 3 shows the result of applying the gradient method to minimize this functional for the cost function *ε* (*s*) = *s*, under continuous policies *s*(*t*) of the form (4) satisfying the critical care capacity constraint (2). The plot shows both the critical care occupancy 𝒞(*t*) and the physical-distancing measure *s*(*t*) between times *t*_0_ and *t*_*f*_ = *t*_0_ + 2 × 365. The aggregate cost of the optimal policy is equal to the economic cost of sustaining a full lockdown for 294 days. As anticipated, the optimal policy found by the computer is very complicated. Notice that the critical care occupancy grows quickly towards the end of the plot. The reason for this is that we asked the computer to minimize the total time in lockdown over a fixed period of two years, which is exactly what it did: it minimized the physical-distancing measures in the first two years with complete disregard for what could happen next. To overcome such a pathological case, one can introduce additional terms into Eq. (7) that would constrain the final slope of the curve 𝒞 (*t*), to make it less steep, or even decreasing.

**FIG. 3:**
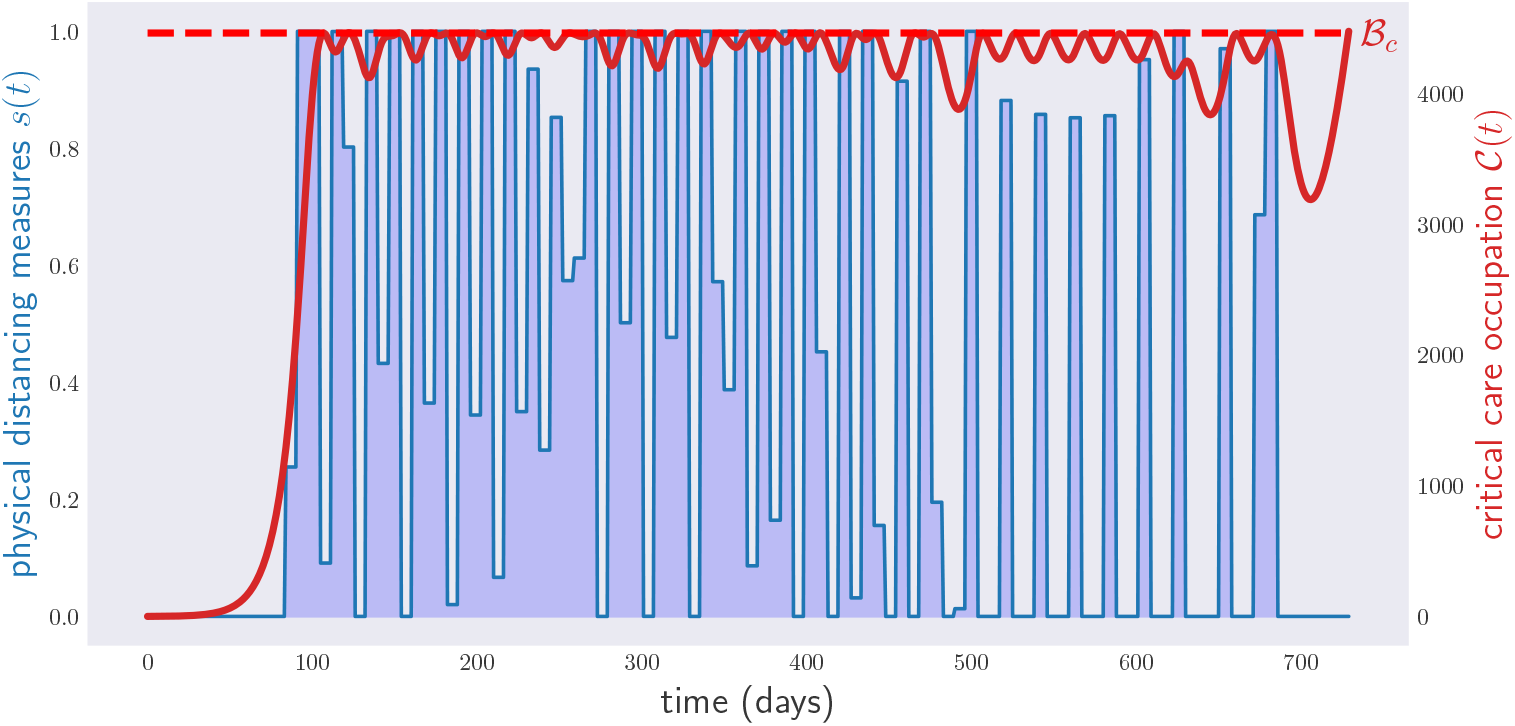
Occupation of critical care beds (red) and physical-distancing measures (blue) for a period of two years. The optimization has been performed over continuous weekly policies, 104 continuous parameters, i.e., any value of physical-distancing measure *s* between 0 and 1 is accepted. The algorithm, however, tends to prefer 0*/*1 configurations, i.e., full lockdown or no lockdown in most instances.

**FIG. 4:**
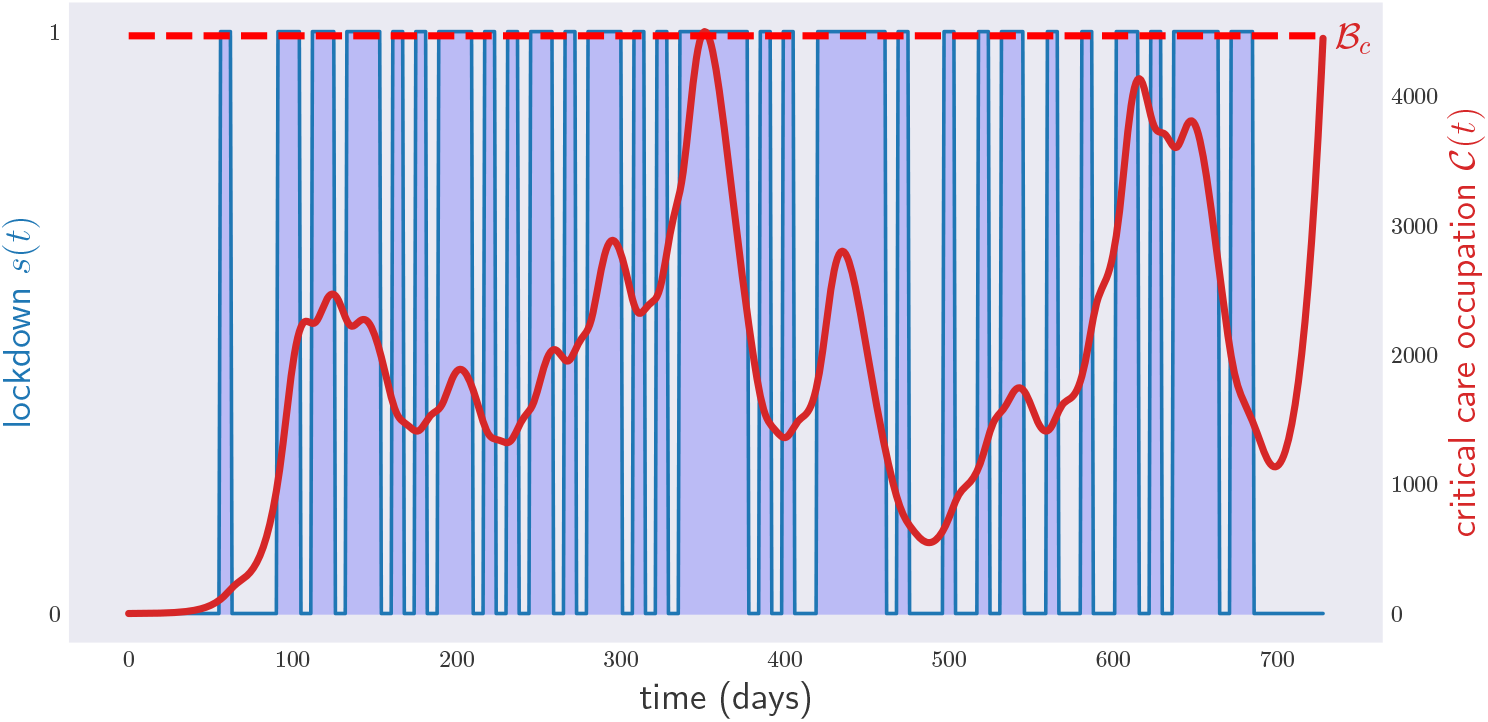
Occupation of critical care beds (red) and lockdown (blue) for a period of two years. The plot shows the result of the optimization over probabilistic policies via gradient descent over a period of two years.

In some circumstances, the only distancing measures considered by governments are discrete: lockdown on or off as in Eq. (1). These kinds of optimizations over discrete variables cannot be carried out directly with the gradient method. In Appendix D, we propose two different ways to tackle this problem.

One way consists of using stochastic gradient descent to obtain an optimal non-deterministic weekly policy that is later “binned” to arrive at a deterministic policy for disease control, see Appendix D. The results of this first method are shown in Figure 4. This time the critical care occupancy curve touches the critical care capacity just after the end of year 1. The reason for this is that we demanded lockdowns to last exactly one week: had we allowed the government to declare a lockdown on any day of the week, the computer would have found a tighter solution, with every peak of the red curve touching the dashed line. Even under this discrete weekly simplification, the solution found by the computer is non-trivial: it requires the government to declare a lockdown 27 *times*. The total length of the lockdown in the course of two years is 371 days.

Our second discretization method consists in parametrizing the disease control policy through the specific times where lockdown is declared. In other words, ***µ*** = (*t*_1_, *t*_2_, …, *t*_2*N*_), and lockdown is assumed to take place within the time intervals [*t*_1_, *t*_2_], [*t*_3_, *t*_4_], In this parametrization, lockdowns can be declared or lifted at arbitrary times within [*t*_0_, *t*_*f*_], and not only on Mondays, like in the first method. This second discretization method has the advantage of allowing one to set the maximum number *N* of lockdowns throughout the period [*t*_0_, *t*_*f*_]. For *N* = 9, the corresponding critical care occupation and lockdown graphs are shown in Figure 5. The total length of the lockdown is 338 days.

**FIG. 5:**
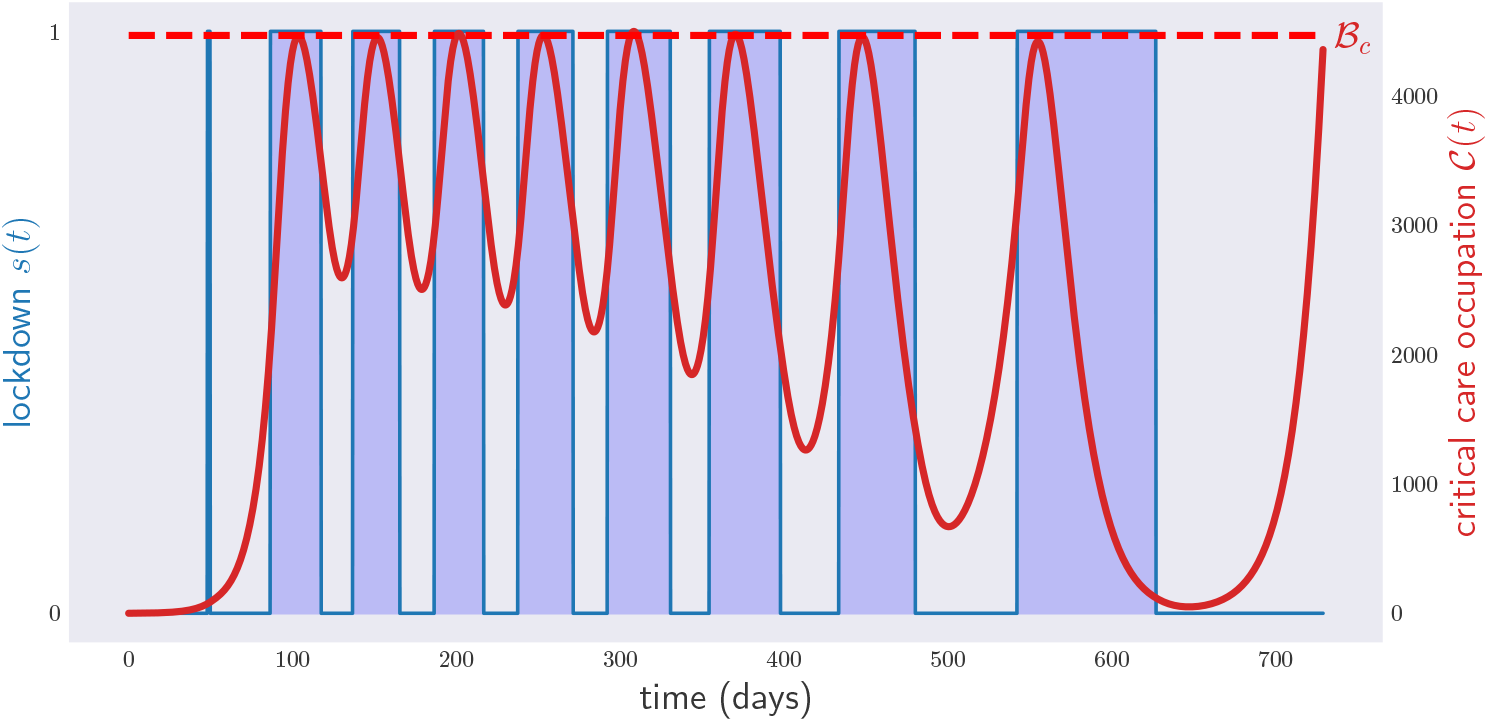
Occupation of critical care beds (red) and lockdown measures (blue) for a period of two years. Optimization over deterministic policies with arbitrary initial and final times for each lockdown period. A total of 9 lockdown periods has been fixed prior to the optimization. Notice that the first lockdown period, around day 60, has been basically removed by the optimization procedure.

### B. Objective function 2: achieving herd immunity

One problem with the policies above is that they require one to implement lockdown measures over long periods of time. Another strategy to fight COVID-19 consists of steering the population towards herd immunity, without violating the constraint on the critical care capacity. This is captured by using a functional of the form

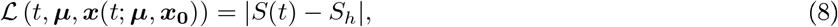

in Eq. (6), where *S* (as depicted in Fig. 1) is the component of ***x*** that denotes the proportion of individuals in the population who are susceptible to the disease. 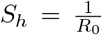 is the proportion of susceptible individuals required for herd immunity to be guaranteed; thus, once *S*(*t*) *< S*_*h*_ although people will continue to become infected, the natural evolution of the disease will be such that the rate and number of infected quickly dies out.

Fig. 6 illustrates the results of optimizing the objective function in Eq. (8) for continuous physical-distancing measures. The number of susceptible individuals reaches *S*_*h*_ on day 864, after which the disease can be considered extinct.

**FIG. 6:**
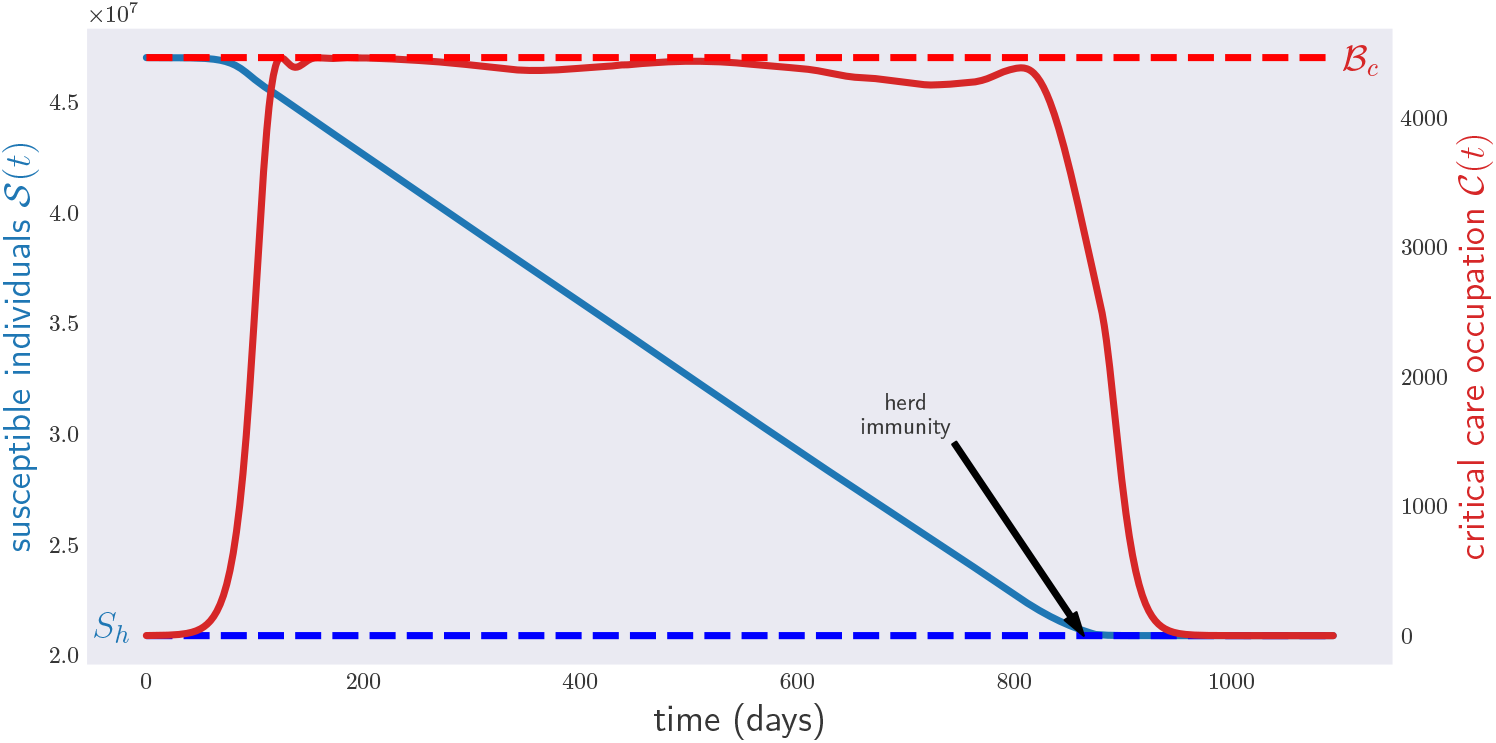
Occupation of critical care beds (red) and population of susceptible individuals (blue) for a period of three years. The blue line corresponds to the level of susceptibles guaranteeing herd immunity over the whole year.

Notwithstanding the unprecedented velocity in the development of a COVID-19 vaccine, with many pharmaceutical companies and research institutions that already reached advanced stages in clinical trials on humans, as of now, no vaccine has completed all testing phases before being approved [28]. One may wonder what will happen if the situation were different with a possible vaccine not available in a time span of months.

A possible policy to reach faster herd immunity, while not saturating the critical care capacity, could be to deliberately infect a fraction of the population at the same time as imposing physical distance measures. It is important to remark that, even allowing this procedure on a voluntary basis among the population which are at the lowest health risk (e.g., young adults in good health), this practice poses high ethical concerns; as such, we remark that we do *not endorse applying such “infection policies;; on a human population*. We just consider them in this paper for the academic purpose of exploring the power of the gradient method to devise complex policies for disease control. In fact, the results of our simulations indicate that the advantage with respect to simple physical distancing policies are minimal (cf. Fig. 7).

**FIG. 7:**
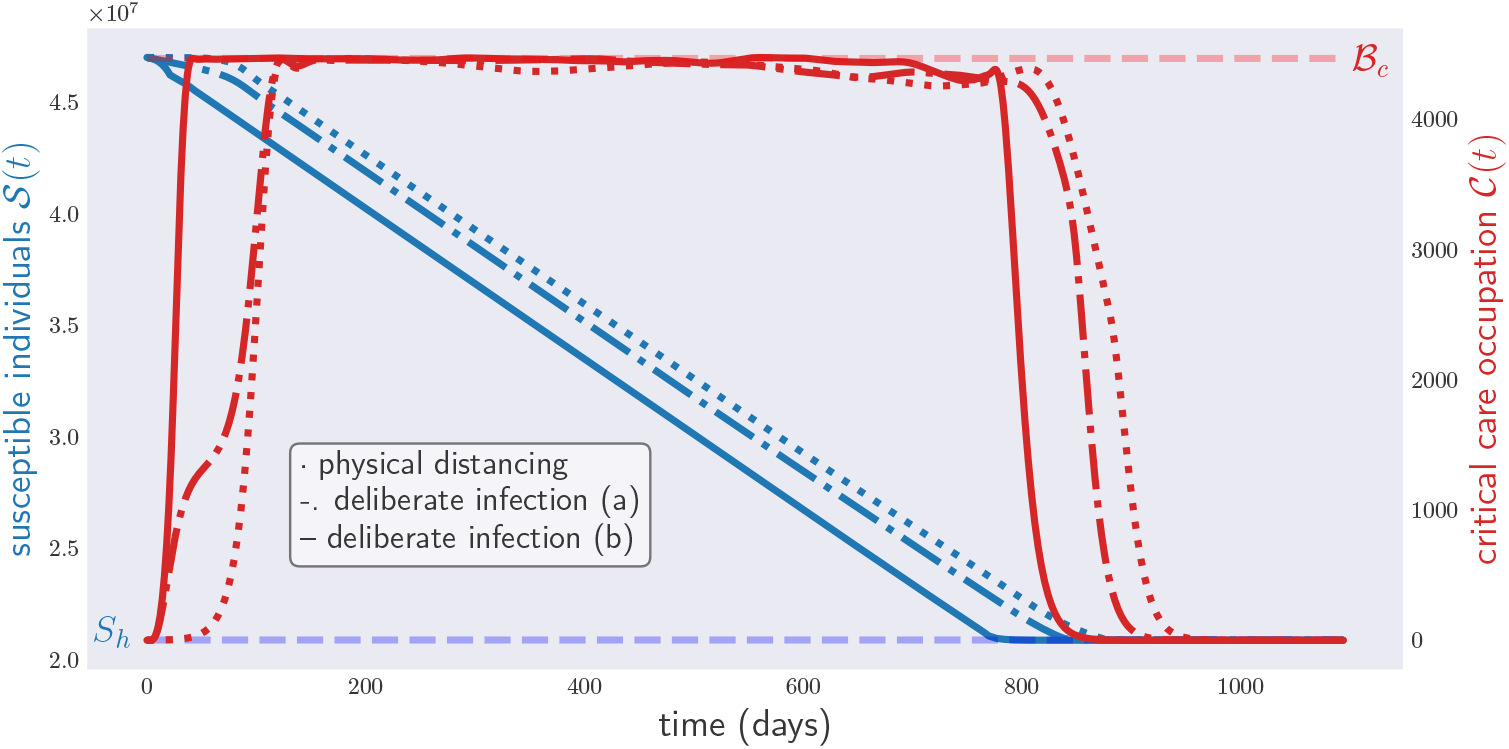
Occupation of critical care beds (red) and population of susceptible individuals (blue) for a period of three years under different policies for disease control. The dotted lines represent the effects a physical distancing measures-only policy (dotted lines), infection policy (a) (dashed-dotted lines) and infection policy (b) (solid line).

That being said, consider the following model of such an *unethical policy*. Let *λ*(*t*) be the rate at which the government intentionally infects the general population on day *t*. Then, a lockdown-infection policy is identified with the functions *s*(*t*) and *λ*(*t*). As with *s*, one can parametrize the function *λ* to change once per week. Denoting by Λ the maximum rate of this deliberate infection, a suitable parametrization for *λ*(*t*) is

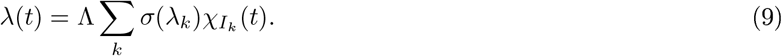

The disease model used to predict the impact of this policy will depend on how the government decides to deal with those infected: (a) the infected are quarantined until they overcome the disease; (b) the infected are allowed to mingle with the general population. In either compartmental model, one can compute the functions ***x***(*t*; ***µ, x***_**0**_), with 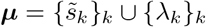. We are interested in minimizing the functional given by (8) for the problem in Eq. (6).

The results for both quarantined/non-quarantined policies for 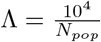 are shown in Figure 7, where we included the curves of the no-infection policy for comparison. As can be seen, the unethical policy of intentional infection brings the population to herd immunity, and thus the disease eventually to extinction, just slightly earlier than the optimal policy based on physical-distancing measures only.

Note how tight the constraint on critical care bed occupation becomes for policy (b). Figure 8 shows the strategy achieving this result. As in the previous cases, the optimal policy is extremely complicated in terms of physical distancing measures. The infection policy is, however, very simple: except at the beginning of the outbreak and after the disease’s extinction, infection occurs at the maximum rate. Notice also that, once the disease is extinct, the computer recommends a permanent lockdown, the reason being that the objective function in Eq. (8), contrary to Eq. (7), does not penalize the overzealous use of physical distancing measures.

**FIG. 8:**
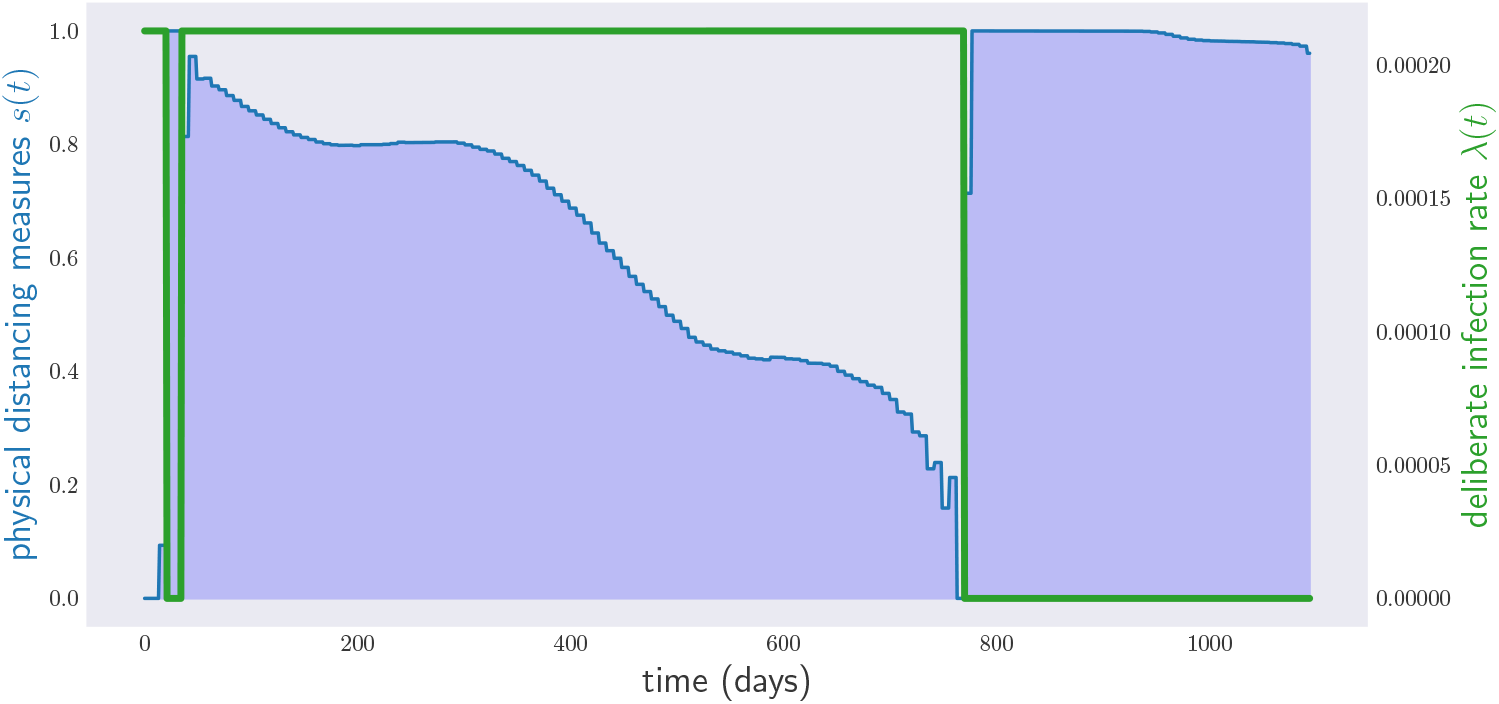
Government infection (green) and physical distancing measures (blue) under infection policy (b). The algorithm recommend an almost constant infection rate, except for a brief initial period, which goes to zero after reaching herd immunity. Notice that, since we are minimizing the distance from the herd immunity threshold (cf. Eq (8)), the algorithm does not care about minimizing physical distancing measures. This is the reason why after reaching herd immunity, a full lockdown is suggested. This type of behavior of the algorithm can be easily corrected by hand after the solution is obtained.

## IV. MODELS VS. REALITY

In practice, the predictions of any mathematical model for a physical system will not be perfect for a number of reasons. First, basic parameters of the model, such as the transmission rate or the initial occupation ***x***_**0**_, are only known up to approximations. Even if reality were exactly described by a particular mathematical model, small errors in such parameters would accumulate in the long run, making long-term predictions unreliable. Second, reality is never exactly described by mathematical models: on the contrary, any tractable disease model is, at best, a rough approximation to reality. Consequently, even the most successful disease models in the market cease to deliver solid predictions beyond 4 weeks [15].

These considerations make us question how practical a two-year disease control policy really is. Consider the policy depicted in Figure 3, which was obtained by applying the gradient method to the SEIR model in [14]. Here, the model parameters ***ν*** correspond to the average values 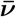 of the parameter ranges in Table A of Appendix A. In reality of course, the values of the parameters are never all equal to their averages, so we proceed to generate sets of parameters with some fluctuations. Let Δ***ν*** be the vector with entries given by the difference between the upper and lower bounds of all the entries of the table, and suppose that the actual parameters of “reality” are unknown and uniformly distributed in the region of values 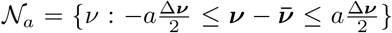, where *a* can be interpreted as the amount of noise or uncertainty. How robust is a policy to uncertainty in the initial parameters?

Fig. 9 shows the result of generating 1000 independent parameter samples from the region 𝒩_0.05_, corresponding to a 5% uncertainty, with respect to the given interval of values, and running the corresponding models for the optimal physical distancing policy in Fig. 3. As one can see, for some sampled values of parameters, the critical care capacity of the healthcare system is exceeded. This is not surprising, since the policy depicted in Fig. 3 was devised to perform well under the assumption that 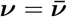 and not ***ν*** *∈* **𝒩** _0.05_, i.e., for the model parameters corresponding to their average value.

**FIG. 9:**
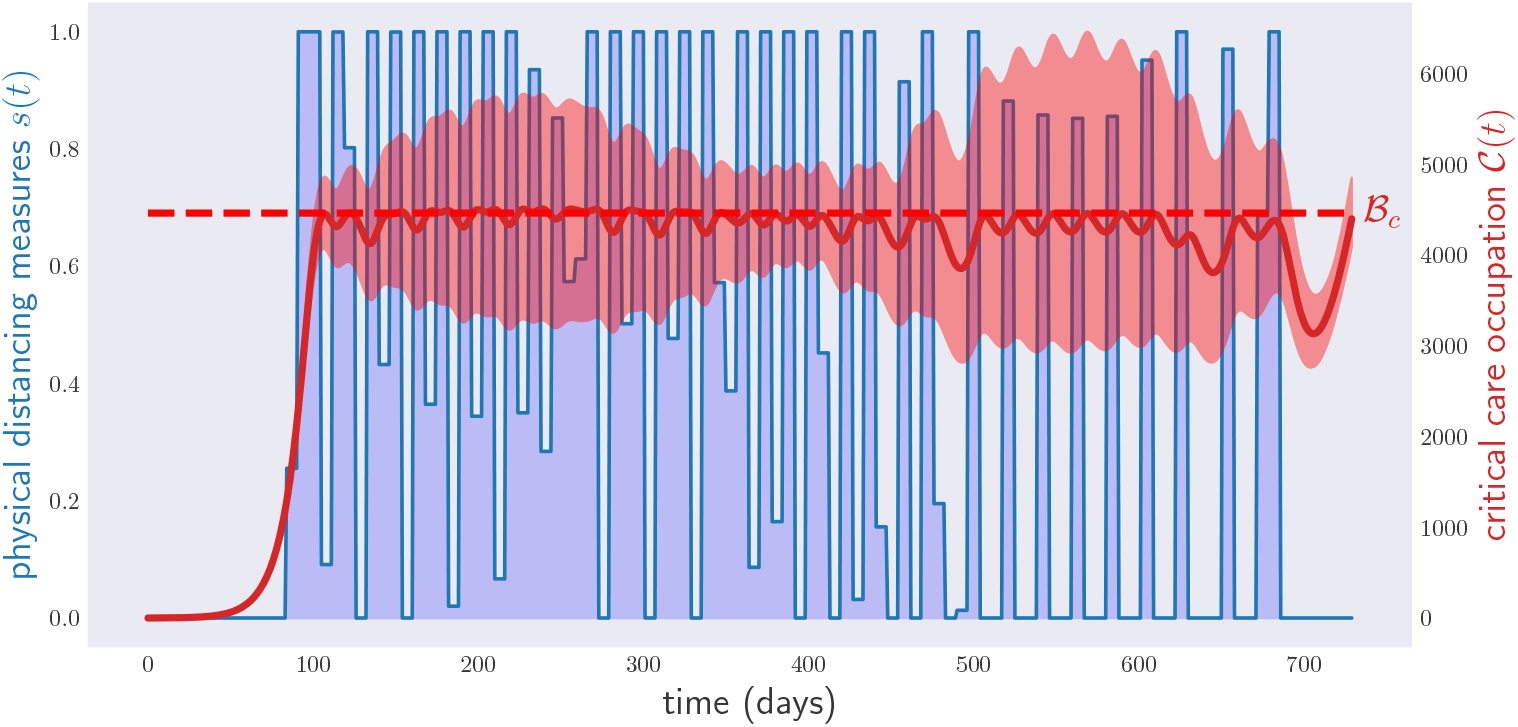
Occupation of critical care beds (red) and (unoptimized) physical distancing measures (blue) for a period of two years under random parameters in 𝒩_0.05_. The region in red is obtained by sampling 1000 times from the region of model parameters 𝒩_0.05_ and evolving the corresponding models with the physical distancing policy optimized over the model with average-value parameters (as in Fig. 3). More precisely, the red region is the one delimited by the minimum and the maximum critical care occupation for all the 1000 models, at each time. The red line represents the average critical care occupation in all those simulations.

In order to tame the behavior in the plot in Fig. 9, one can employ a variant of the gradient method called *stochastic gradient descent* (see Appendix B). This allows one to optimize over long-term policies without violating the constraints of the problem for a range of values for the parameters of the model as well as the initial conditions ***x*_0_**^2^. Fig. 10 shows a lockdown policy minimizing the physical distancing measures under the condition that constraint (2) holds for different values of *ν ∈* 𝒩 _0.05_, i.e., the critical care capacity is not exceeded. This time, the violation of condition (2) is neither so extreme nor so frequent. This comes, however, at the cost of enforcing physical distancing measures with a cost equivalent to 331 days of lockdown. Repeating the optimization for 𝒩_0.25_, Fig. 11, we see that the critical care capacity is rarely surpassed. However, this time the total cost is equivalent to 414 days of lockdown.

**FIG. 10:**
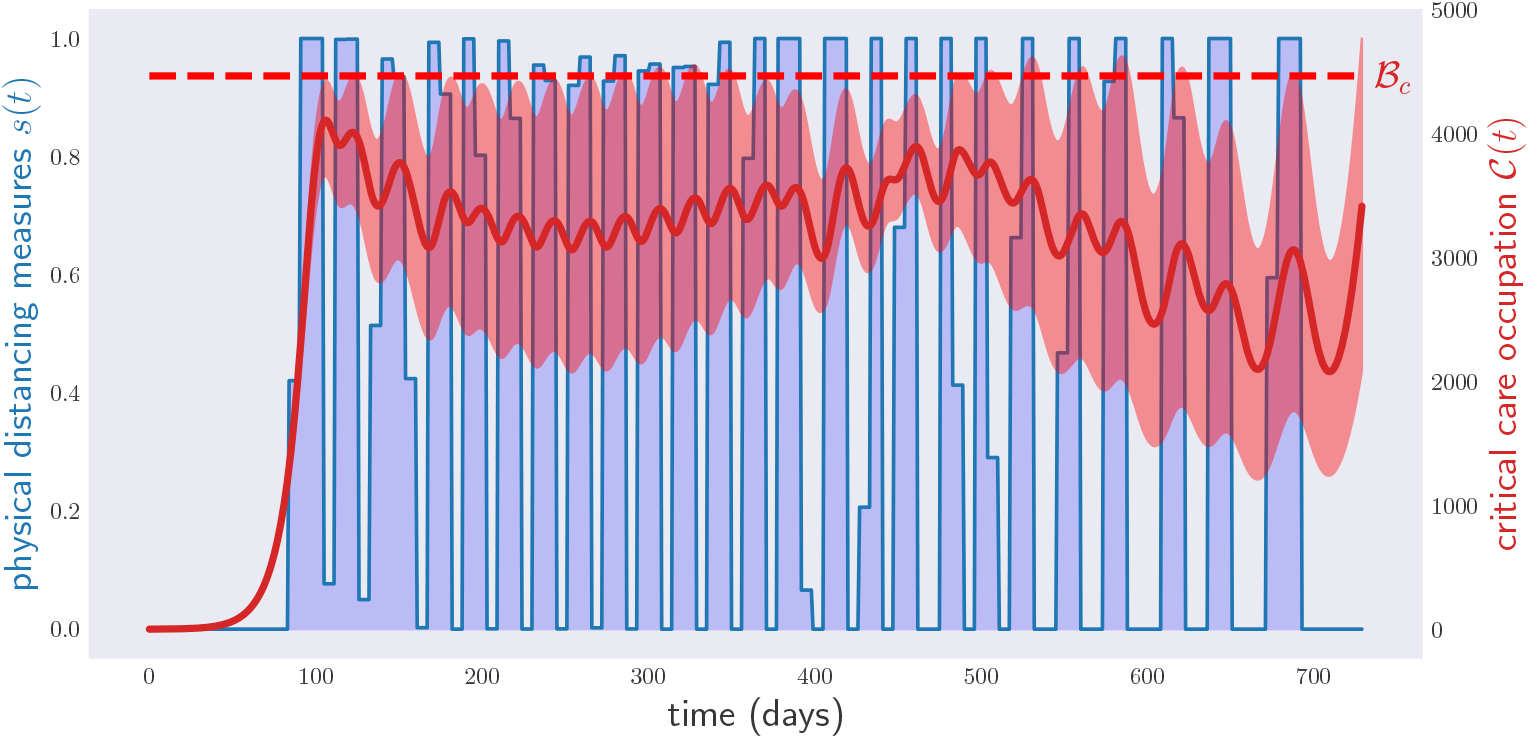
Occupation of critical care beds (red) and (optimized) physical distancing measures (blue) for a period of two years under random parameters in 𝒩_0.05_. The disease control policy was optimized to respect condition (2) over the whole range of parameters 𝒩_0.05_. As in Fig. 9, the region in red depicts again the range of critical care occupations observed in a sample of 1000 model parameters in 𝒩_0.05_, and the red line the average critical care occupation.

**FIG. 11:**
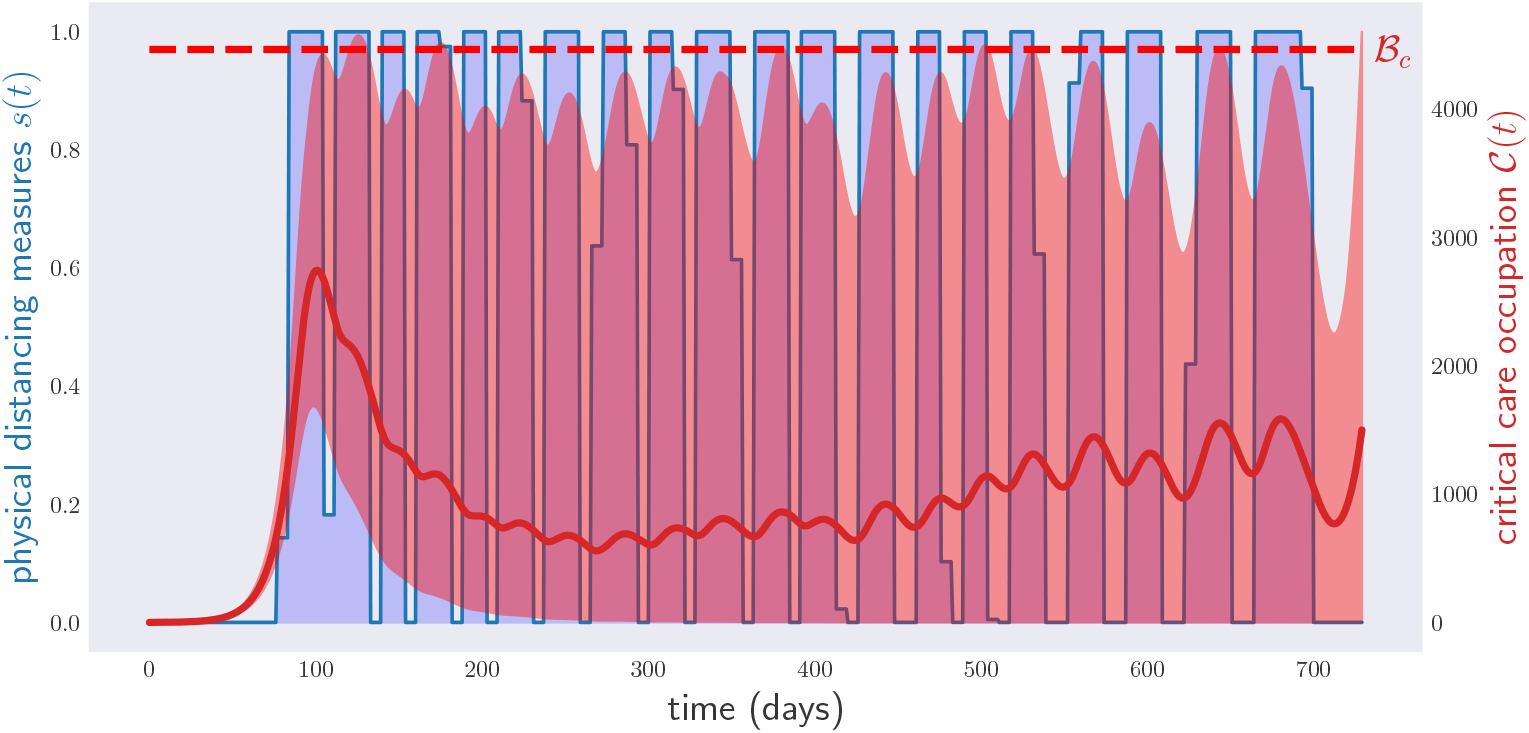
Occupation of critical care beds (red) and (optimized) physical distancing measures (blue) for a period of two years under random parameters in 𝒩_0.25_. The disease control policy was optimized to respect condition (2) over the whole range of parameters 𝒩_0.05_. As in Fig. 9, the region in red depicts again the range of critical care occupations observed in a sample of 1000 model parameters in 𝒩_0.25_, and the red line the average critical care occupation.

This result is what one would have expected. As time goes by, the predictions of the disease model for different values of ***ν*** diverge: any policy which aims to satisfy constraint (2) for large ranges of these parameters will necessarily require extensive physical distancing measures.

In practical policy-making, graphs such as Figs. 10 and 11 should not be understood to represent the actual physical distancing policy, but rather to provide a provisional *policy plan*. A policy plan gives a recommendation for action for the immediate future, given the current knowledge of the disease. In Fig. 10, the policy plan is advising not to declare physical distancing measures in the first weeks. That is the measure that the government should adopt then. After a first time period, say four weeks, more data will have been gathered: this will allow us to obtain a better estimate of the parameters ***ν***, and then re-run the models for another two years ahead. The measure to enforce should then be whatever the new policy plan recommends for the following four-week time period. The process is then repeated.

To test how this idea would perform in practice, we consider a scenario where the parameters defining the disease model are unknown, but the region in parameter space in which they live shrinks every month (to be precise, we used a 28-day period, corresponding to four weeks). That is, at month *k*, the government is informed that the parameters ***ν*** satisfy 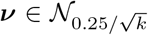. Every four weeks, the policy is recalculated to minimize the physical distancing measures for the rest of the two years ahead, using the range 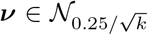. The final curves for the critical care occupation and the physical distancing measures are shown in Fig. 12 for ***ν***, in a sequence of shrinking regions 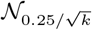, for each month *k* (red region) and in the case of a fixed uncertainty region corresponding to the last month, i.e., 𝒩 _0.049_ (inner dark blue region). The total cost of the physical distancing measures is equivalent to 358 lockdown days. This has to be compared with the cost of 414 days predicted by the initial policy plan under the assumption *ν ∈* 𝒩_0.25_.

**FIG. 12:**
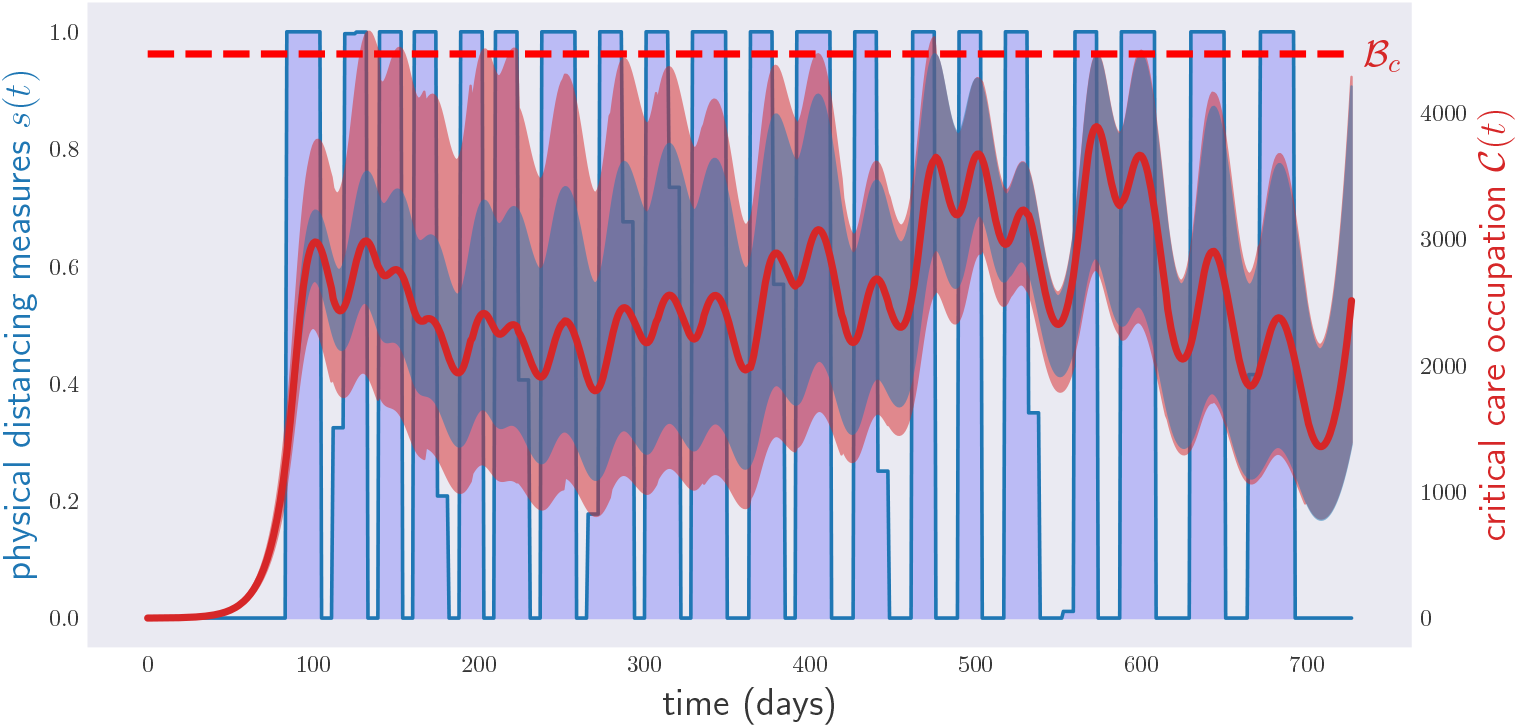
Occupation of critical care beds (red) and physical distancing measures (blue) for a period of two years under random parameters and monthly noise decrease. The disease control policy was optimized to respect the condition in Eq. (2), i.e., critical care capacity not exceeded, starting w<u>i</u>th the parameter region 𝒩_0.25_ and with a monthly noise decrease of 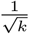, i.e., in the *k*-th month the noise is equal to 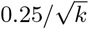. The region in red depicts the range of critical care occupations observed in a sample of 1000 model parameters in a sequence of shrinking regions 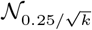; more precisely, for each month *k* the red region is obtained by evolving, from the initial time to month *k* (included), 1000 different models with parameters sampled from the region 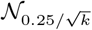. The final plot is obtained by joining the plots for each month *k*. The inner region in dark blue depicts the range of critical care occupations corresponding to the uncertainty in the final month, i.e., obtained with 1000 models with parameters sampled in 𝒩_0.049_. The red line represents the average critical care occupation, obtained by joining the average of the simulations with decreased uncertainty for each month *k*. Despite the initial uncertainty on the parameters *ν* of the disease model, the final lockdown time is much lower, due to monthly revisions of the original policy plan.

In principle, one could further decrease the total planned cost by devising *adaptive policy plans*, where the measure to be taken at each moment depends not only on the current time *t*, but also on the past history of physical distancing measures and their observed effects. In fact, we tried optimizing over adaptive policies described by a continuous version of a neural network architecture known as Long Short-Term Memory (LSTM) [29]. In all our numerical experiments, such simple LSTM architectures could not improve the performance of non-adaptive strategies, but this could be due to ineffective training on our side.

## V. CONCLUSION

In this paper, we have applied standard tools from optimization theory and machine learning to identify optimal disease control policies, given an epidemiological model. This is in stark contrast to standard practice in mathematical epidemiology, where human intuition is used to narrow down the considered set of policies to a uni-parametric family. We saw that the optimal solutions found by our algorithms are highly counter-intuitive, and thus unlikely to be identified by a human. This supports the idea that policies for disease control should be based on a combination of both human expertise and machine learning.

Compared to previous approaches that tried to identify suitable disease control policies through optimal control theory, our framework allows one to devise policies that satisfy arbitrary constraints under arbitrary uncertainties in the initial conditions and model parameters that determine the disease’s dynamics. Our methods, in addition, allow one to optimize over discrete policies, as well as policies that can just vary at certain fixed times.

To illustrate our ideas, we studied a scenario in which a computer is tasked with outputting the minimal amount of physical distancing measures for an epidemic in a hypothetical country, in such a way as to never exceed the critical care bed capacity. We looked at situations in which these measures were continuous (recommendations on the interval [0, 1]) as well as discrete (either 0 or 1) - a lockdown that is off or on, respectively for periods of 2 years. We experimented with measures which are just allowed to change weekly, as well as those in which there is a maximum number of lockdowns that is allowed to be declared. When considering unethical interventions, in which parts of the population are deliberately infected, we found that there is little difference in the time for the disease to become extinct, compared with the natural evolution of the disease.

We examined the problems that one may encounter when applying these techniques to scenarios where the model parameters are not known with high accuracy, which led us to propose practical policy plans which must be continually revisited, to account for our ever-changing and ever-growing knowledge in an epidemic. We tested the viability of this approach by simulating a scenario where the uncertainty on the disease model parameters decreases with time. As expected, the final policy implemented was safe for the final range of parameters and required considerably less physical distancing measures than the initial policy plan hinted.

In this last regard, an interesting problem for future research is how to devise adaptive policy plans for disease control, where the actual measure at each time depends on the whole history of disease indicators accessible to the government. In theory, such plans should predict lower values of the average objective function in scenarios where the model parameters are unknown. In our experience, though, gradient descent alone seems to be unable to beat the non-adaptive score.

Finally, we would like to remark once more that, since the optimization problems we dealt with in this paper are non-convex, the gradient method is not guaranteed to converge to the minimum of the (average) objective function. While conducting this research, in order to convince ourselves that the solutions found by our numerical methods were close to optimal, we had to repeat our optimizations several times, with different initial policies ***µ***^**(0)**^ and learning rates *ϵ*. Such a redundant use of computational resources would have been entirely avoidable if we had had some rough approximation to the exact solution of the problem. Hence we conclude this paper with a challenge for the operations research community: develop mathematical tools which allow one to *lower bound* the solution of minimization problems involving ordinary differential equations.

## Data Availability

Soon we will make our computer codes available.

## Acknowledgments

We thank Luca Gerardo-Giorda and Mario Budroni for useful discussions. C.B. and Y.G. acknowledge funding from the Austrian Science Fund (FWF) through the Zukunftskolleg ZK03.

## APPENDIX A: MODELS FOR THE SPREAD OF COVID-19

In all our numerical simulations, we will assume that the dynamics of the COVID-19 are well approximated by a compartmental model of the SEIR type. When the government policy reduces to enforcing physical distance measures, we will adopt a simplified version of the model used in [14]. This model divides those infected with COVID-19 into three different compartments: *I*_*R*_ or those who recover by themselves from the disease; *I*_*H*_, those who require hospitalization but do not enter a critical care unit; and *I*_*C*_, those who are both hospitalized and visit a critical care unit before recovery. The dynamics of the model are governed by the system of ordinary differential equations below, see the diagram in Figure 13:

**FIG. 13:**
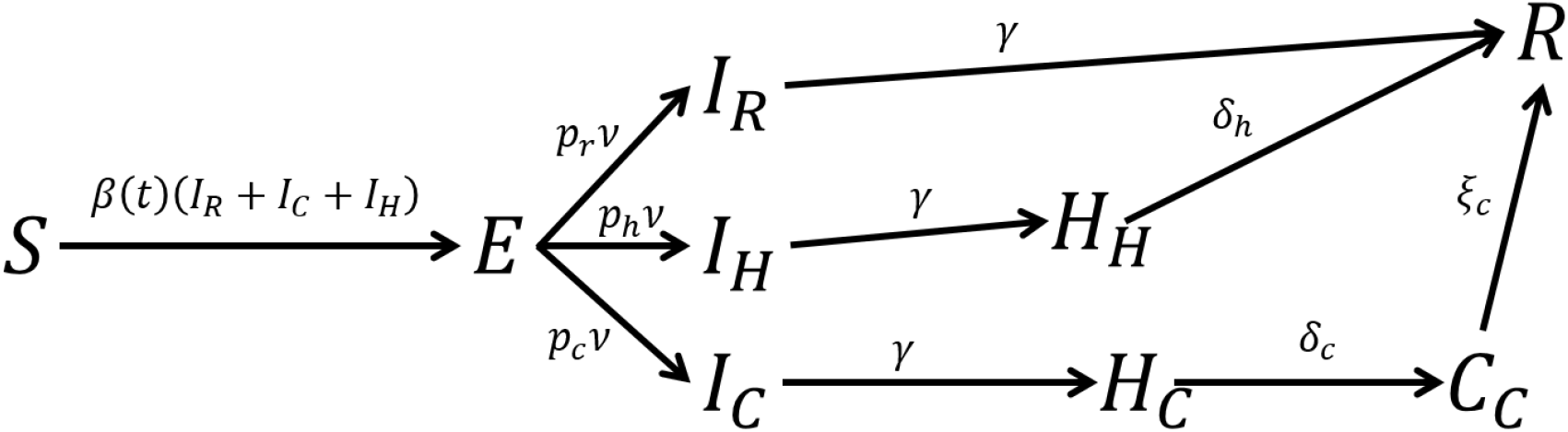
COVID-19 model for disease policies based on physical distance measures.

**FIG. 14:**
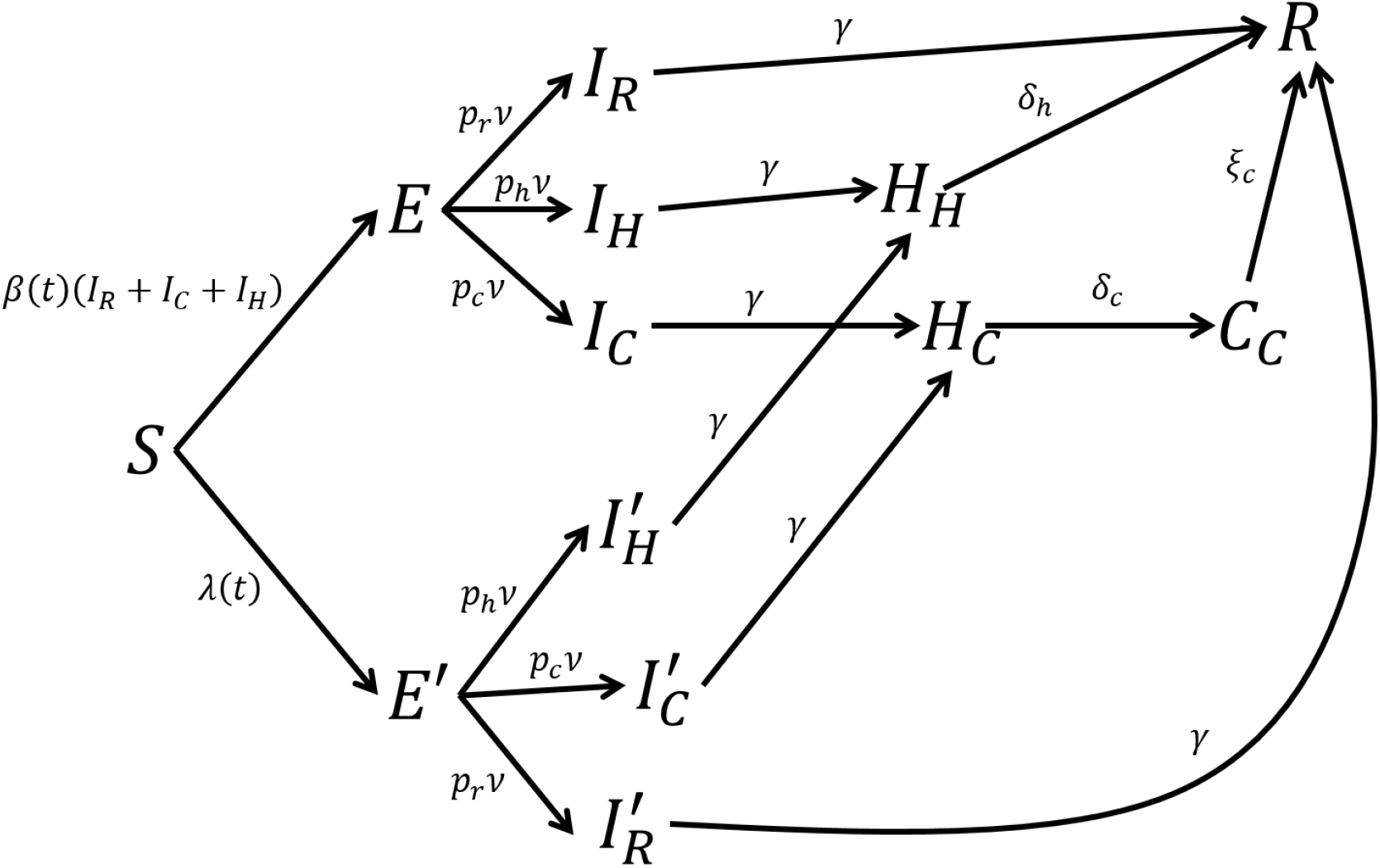
COVID-19 model for disease policies based on physical distance measures and deliberate inoculation.

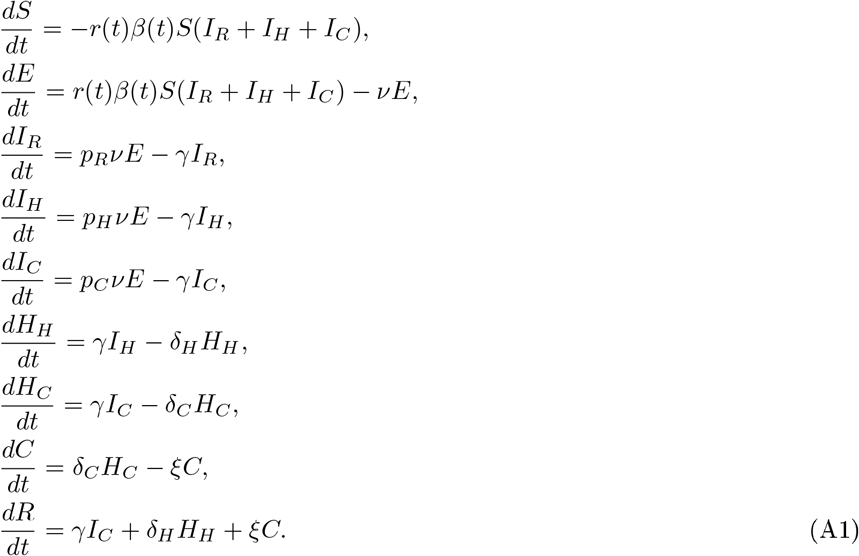

Here

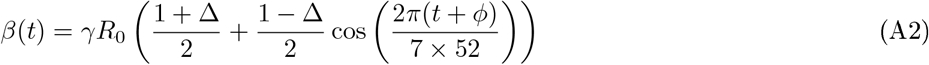

denotes the virus’ transmitivity, that is subject to seasonal variability. 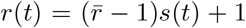 models the effect of a government-mandated lockdown *s*(*t*) *∈* {0, 1}on the virus’ transmission rate. The values of the remaining parameters are taken from [14], and appear in Table A.

**TABLE I:** Parameter ranges for the compartmental model for COVID-19 proposed in [14].

| parameter | value | units |
| --- | --- | --- |
| $\bar{r}$ | $[0, 0.6]$ | none |
| $\Delta$ | $[0.7, 1.0]$ | none |
| $\gamma$ | $1/5$ | days<sup>-1</sup> |
| $\nu$ | $1/4.6$ | days<sup>-1</sup> |
| $p_R$ | 0.9596 | none |
| $p_H$ | 0.0308 | none |
| $p_C$ | 0.0132 | none |
| $\delta_H$ | $1/8$ | days<sup>-1</sup> |
| $\delta_C$ | $1/6$ | days<sup>-1</sup> |
| $\xi$ | $1/10$ | days<sup>-1</sup> |
| $\phi$ | $-7 \times 3.8$ | days |
| $R_0$ | $[2, 2.5]$ | None |

If the government has the option of deliberately infecting random samples of the population with COVID-19, then the model gets slightly more complicated. If those infected are quarantined until they overcome the disease, the new model, depicted in Figure 14, is described by the system of equations:

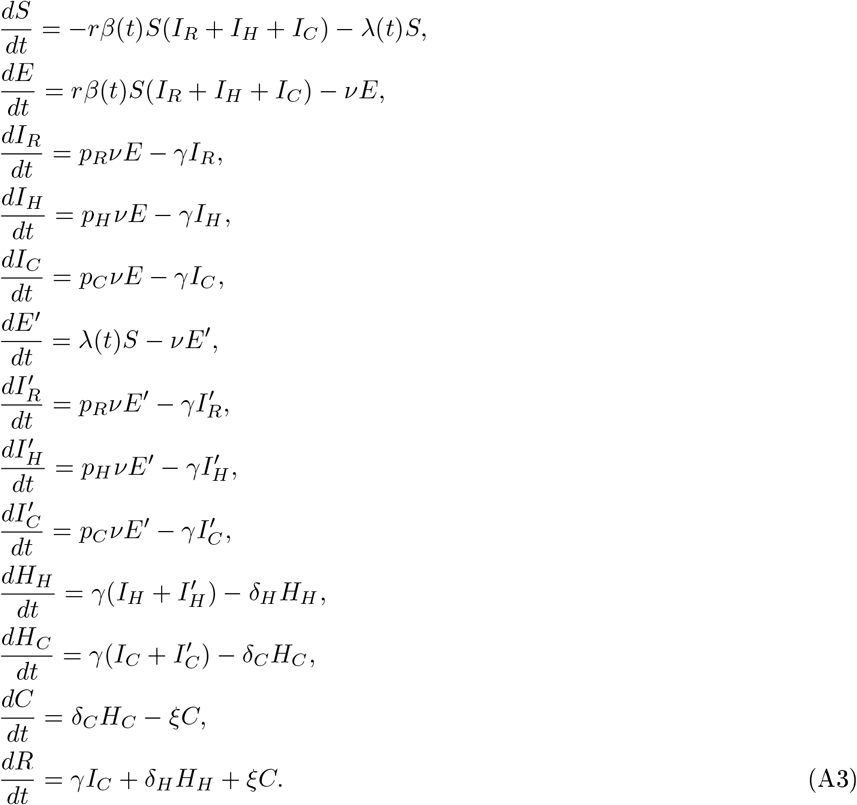

If, on the contrary, the government allows those deliberately infected to mingle with the general population, the equations above simplify considerably: it suffices to replace all instances of the term *rβ*(*t*)*S*(*I*_*R*_ + *I*_*H*_ + *I*_*C*_) in eqs. (A1) by *rβ*(*t*)*S*(*I*_*R*_ + *I*_*H*_ + *I*_*C*_) + *λ*(*t*)*S*.

In all our numerical simulations, we take the total population to be 47 million; the critical care bed capacity per inhabitant *C*_*c*_ is also taken to be 9.5 *×* 10^−5^. We assume that the government starts its intervention on day *t*_0_ = 60, 30 days after the outbreak of the disease. We model the disease outbreak by assuming that, at time *t*_*out*_ = 30, there are 10 individuals in compartment *E*. By default, the values of the disease parameters are taken to be the arithmetic means of the intervals shown in Table A. We assume that the government can deliberately infect 10^4^ individuals per day. This sets a value for Λ of 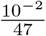.

## APPENDIX B THE GRADIENT METHOD

Given functions *f, g*_*i*_ : ℝ^*n*^*→*ℝ, for *i* = 1, …, *K*, consider the following optimization problem:

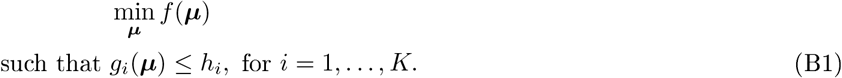

Each of the conditions *g*_*i*_(***µ***) *≤ h*_*i*_ is called a *constraint*.

Define *M* = {***µ*** : *g*_*i*_(***µ***) *≤h*_*i*_, *i* = 1, …, *K*}. If *f* is sub-differentiable, a simple heuristic to solve this problem is the projected gradient method [22]. Call ***µ***^*\**^ the solution of the problem. Starting from an initial guess ***µ***^(0)^, the gradient method generates a sequence of values (***µ***^(*k*)^)_*k*_ with the property that lim_*k→∞*_ ***µ***^(*k*)^ = ***µ***^*\**^, provided that *M, f* are, respectively, a convex set and a convex function [22]. The sequence (***µ***^(*k*)^)_*k*_ is generated recursively via the relation

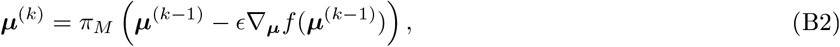

where, for any set *A ∈* ℝ^*n*^, *π*_*A*_(***z***) denotes the point ***y*** *∈ A* that minimizes the Euclidean distance, i.e., min_***y****∈A*_ ∥***y* − *x* ∥**_2_. Unfortunately, in the problems we encounter in the main text, *f* is not convex and, sometimes, neither is *M*. This implies that the sequence output by the projected gradient method is not guaranteed to converge to the solution of the problem, but to a local minimum thereof.

In machine learning, optimization problems with non-convex objective function *f* and *M* = ℝ^*n*^are legion. To solve them, deep learning practitioners typically use variants of the gradient method sketched above. One of these variants, Adam [30], is extensively used to train neural networks.

Adam works as follows. Starting with the null vectors ***m***^**(0)**^, ***v***^**(0)**^ *∈* ℝ^*n*^, vector sequences are generated according to the following iteration rule:

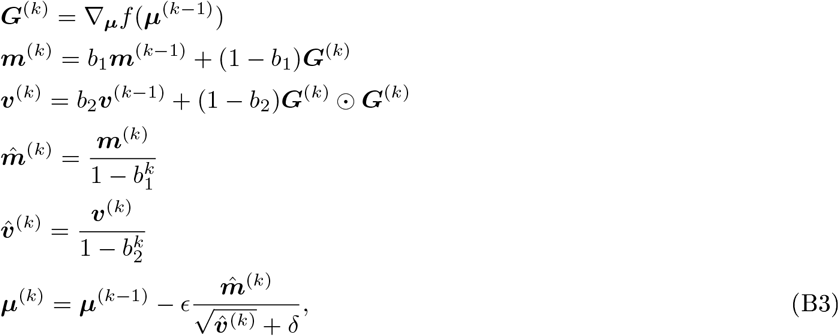

where *G*^(*k*)^ ⊙ *G*^(*k*)^ denotes the vector of the element-wise product; similarly, the fraction and square root in the definition of ***µ***^(*k*)^ are defined element-wise. Recommended values for the free parameters *ϵ, b*_1_, *b*_2_, *δ* are *ϵ* = 0.001, *b*_1_ = 0.9, *b*_2_ = 0.999 and *δ* = 10^−8^ [30].

Like the projected gradient method, Adam is not guaranteed to converge to the optimal solution of the problem. However, provided that the initial conditions and the learning rate *E* are chosen with care, Adam has been observed to typically output a local minimum that is “good enough”.

Note that, by taking *M* = ℝ^*n*^, it is not clear how to enforce that the solution satisfies constraints of the form *g*_*i*_(***µ***) *≤ h*_*i*_. The answer is to include those constraints as penalties in the objective function. That is, rather than minimizing *f*, we apply Adam to minimize the function

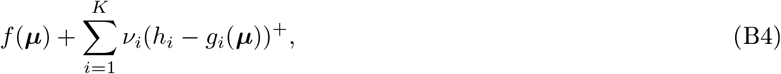

where *ν*_*i*_ ≫ 1 and *z*^+^ denotes the positive part of *z*, i.e., *z*^+^ = *z* for *z >* 0; otherwise, *z*^+^ = 0. For high enough values of *ν*_*i i*_, the solution of the problem will just violate the constraints slightly, i.e., *h*_*i*_ − *g*(***µ***^*\**^) *∈* [− *δ, ∞*), for *δ* ≪ 1. If no violation whatsoever is desired then one can instead optimize over a function of the form

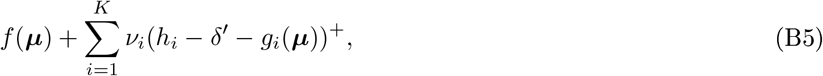

with *δ*^*′*^*>* 0.

In some situations, the objective function *f* will be complicated to the point that computing its exact gradient is an intractable problem. It might be possible, though, to generate a random vector 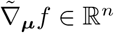with the property

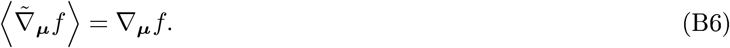

In such a predicament, we can solve the original optimization problem (B1) through stochastic gradient descent methods [23]. Stochastic gradient descent consists in applying the considered gradient method, with the difference that, every time that the method requires the gradient of *f*, we input the random variable 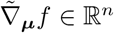 instead. Namely, it suffices to replace 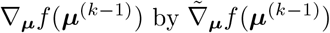 in the iterative equations (B2), (B3). As before, if both *M* and *f* are convex, stochastic gradient descent methods are guaranteed to converge to the optimal solution of problem (B1) [23].

## APPENDIX C: OPTIMIZATION OVER CONTINUOUS POLICIES FOR DISEASE CONTROL

Our starting point is an ordinary differential equation of the form

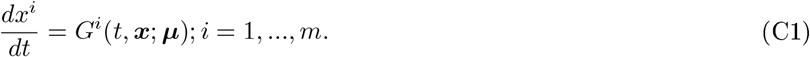

The entries of vector *x* represent the occupations of the different compartments of a disease model^3^. ***µ*** *∈* R^*n*^represents a parametrization of the effects of a given policy. In the examples of the main text, 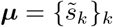 or 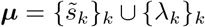. Call 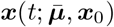 the solution of Eq. (C1) with initial conditions ***x***(0) = ***x***_0_ and 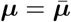.

Given the set *M ⊂ℝ*^*n*^, we consider the problem of finding the parameters ***µ***^*\**^ *∈ M* such that ***x***(*t*; ***µ***^*\**^, ***x***_0_) minimizes a given functional *A*. This functional defines how we wish to control the disease and what for: it might represent the number and duration of lockdown, etc. For the time being, let us assume this functional to be of the form

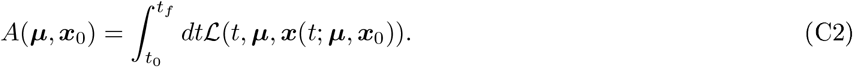

Note that we are assuming to know the initial conditions ***x***_0_ with precision. We will relax this requirement by the end of the section.

From the discussion in section B, functional (C2) might also contain constraints which we wish the solution ***x***(*t*; ***µ***^*\**^, ***x***_0_) to satisfy. For instance, if we want Eq. (2) to hold, then one of the terms in ℒ(*t*, ***µ, x***(*t*; ***µ, x***_0_)) could be

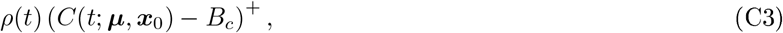

with *ρ*(*t*) ≫1. In the optimizations presented in the paper, we did just this, with 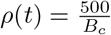 (figures 12, 11), 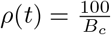 (figure 10) or 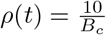 (rest of the figures).

As we saw in section B, optimizations over a large number of parameters ***µ*** *∈* ℝ^n^ are usually conducted via gradient descent methods, such as Adam [30]. Such methods require us to compute *∇*_***µ***_*A*. For functionals of the form (C2),

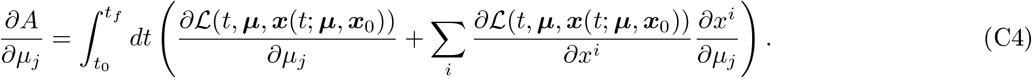

The next question is thus how to compute the derivatives 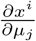. To this aim, define the variables 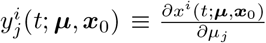. Differentiating equation (C1) by *µ*_*j*_, we have that

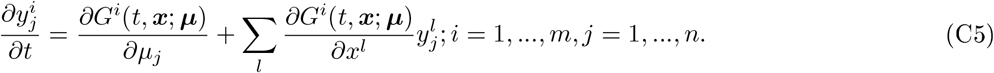

In order to obtain 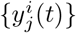 for each time *t*, it hence suffices to solve the system of coupled differential equations given by (C1), (C5) with initial conditions 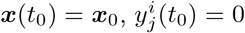. This can be achieved numerically through several different methods, depending on the desired accuracy. In all our numerical simulations, we used the Euler explicit method [31]. Namely, for *δ >* 0, we regarded time as a discrete variable of the form *t*_*k*_ = *t*_0_+δ_*k*_, for 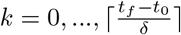.

We obtain the quantities 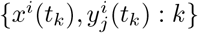 by recursively applying the relations

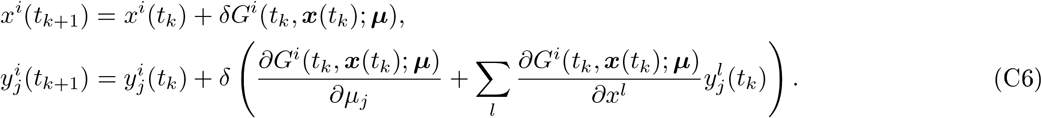

We chose *δ* = 1.0 to generate all our plots, with the exception of Figure 5, where *δ* = 0.1.

In some situations in the main text, our functional *A* is more complicated than (C2). Some parameters ***ζ*** (not policy parameters) regulating the evolution (C1), such as the disease’s basic reproduction number, might be unknown, or perhaps the initial conditions ***x***_0_ are just known within some bounds. In such cases, the problem’s objective function *A* might adopt the form

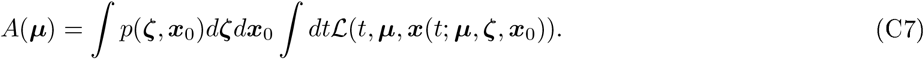

Again, we wish to minimize *A* over *λ ∈ M*. As explained in section B, this can be achieved via stochastic gradient descent methods [22]: all we need is an unbiased estimator 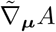 of *∇*_***µ***_*A*. We obtain this estimator by taking **𝒩** independent samples 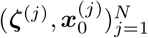 from the measure *p*(***ζ, x***_0_)*d****ζ****d****x***_0_ and using them to compute the quantity

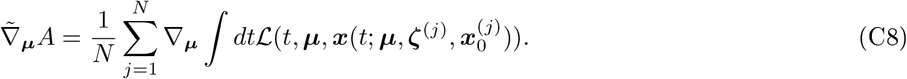

The optimal value of *N* depends on the variance of the specific estimator, and we used different values for different levels of noise, defining the set **𝒩** in the main text.

## APPENDIX D: OPTIMIZATION OVER DISCRETE POLICIES OF DISEASE CONTROL

The above section explains how to conduct optimizations over disease control policies, as long as the parameters ***µ*** defining the policy are allowed to vary all over ℝ^*n*^. Some policies, though, are by their very nature, discrete. For instance, on day *t* we can either declare a lockdown (*s* = 1) or not declare a lockdown (*s* = 0). The effect of the policy measure over the disease’s transmission rate will be to multiply the latter by the amounts 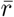 and 1, respectively.

In principle, we could model this situation via a continuous variable *λ ∈* ℝ and write the effect *r* on the transmitivity by means of a piece-wise continuous function of *λ*, e.g.: 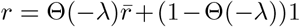, where Θ(*z*) denotes the Heaviside function ^4^.

In that case, however, the gradient method would not work. Note that the Heaviside function has zero derivative everywhere except at 0. In every iteration of Adam, *∇*_*λ*_*A* would be null, and so *λ*^(*k*)^ = *λ*^(0)^ for all *k*.

Optimizing over discrete variables is, in general, a very difficult endeavor: apparently simple problems can be argued not to have an efficient solution [32]. In this paper, we propose a simple heuristic to attack this problem in this particular situation.

Suppose, for the time being, that our lockdown policy were probabilistic, i.e., at each week *k*, we declare a lockdown with probability 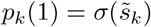; otherwise, with probability 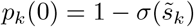, we let the population roam freely. We wish to minimize our average objective function, that is, the expression

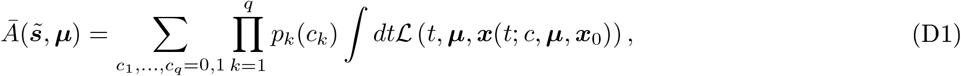

where *c* is the whole vector of weekly lockdowns, and ***µ*** corresponds to the continuous parameters of the policy, e.g.: vaccination rates.

In principle, we could apply gradient descent to minimize (D1). Estimating the exact gradient of the above expression is, however, unrealistic, as it involves summing a number of terms exponential in the number of weeks *q*. Instead, we will produce a random unbiased estimate of the gradient and invoke stochastic gradient descent methods, see Appendix B.

Let us first differentiate Eq. (D1) with respect to the continuous variables ***µ***. The result is

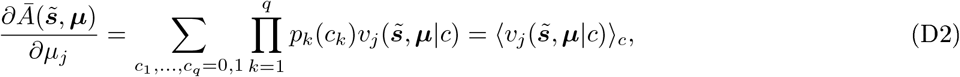

where

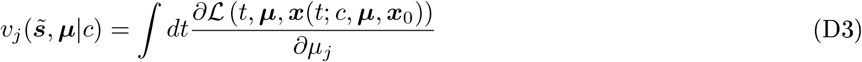

and the components of the random variable *c* 0, 1 ^*q*^ are generated by sequentially sampling from the Bernouilli distributions 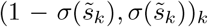. Note that the expression in the integrand of (D3) can be computed using the techniques discussed in Appendix C.

Differentiating Eq. (D1) with respect to 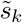 we find that

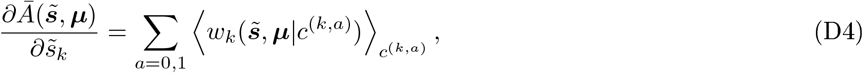

with

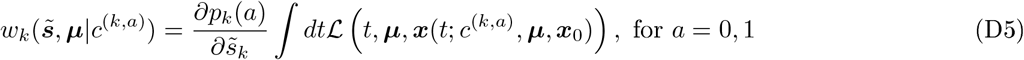

and the average 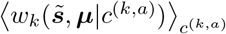 is obtained via sampling over the product of Bernoulli distributions for 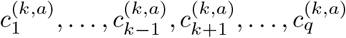 and fixing 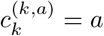.

Putting this all together, we have that the random vectors *v, w* satisfy

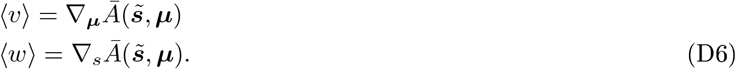

Since both vectors can be sampled efficiently, we can use them (and their averages) to optimize over 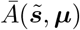 via stochastic gradient descent.

At this point, the reader might object that our original goal was to minimize (C2) over policies with *deterministic* lockdown. Very conveniently, independently of the initial values of *s*, ***µ*** the stochastic gradient method will converge to a policy *p*^*\**^, ***µ***^*\**^ such that the deterministic policy with the same continuous parameters ***µ***^*\**^ and lockdown given by

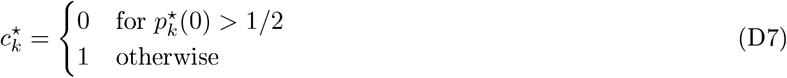

has the same objective value.

Indeed, for *k ∈ {*1, …, *n}*, fix 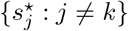. Then,

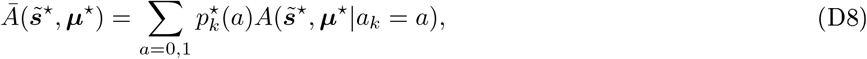

with

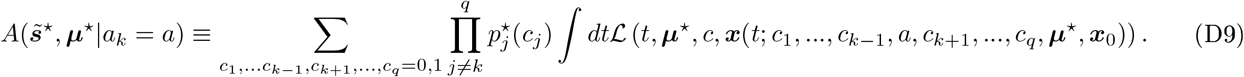

Since *p*^*\**^, ***µ***^*\**^ is a local minimum of 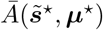, it follows that, either 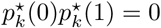 or

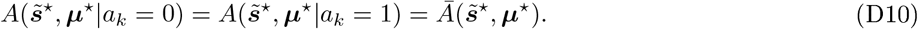

In either case, fixing *c*_*k*_ through the procedure (D7) cannot increase the average value of the objective function. Iterating over *k* = 1, …, *q*, we prove the claim.

Note that this method easily extends to optimizations over discrete adaptive policies. In that case, one can model the probability *p*_*k*_ of lockdown on week *k* as a function of both the current input 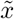 and the cell state ***θ***, i.e., 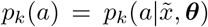. Let us assume that the family of functions 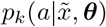 available is rich enough to regard 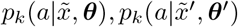 as independent. Then one can argue as before and conclude that a deterministic policy can be inferred from the limiting probabilistic policy generated by stochastic gradient descent.

### 1. Optimization over discrete policies with continuous lockdown times

In this section, we explain how to optimize over lockdown policies of the following form:

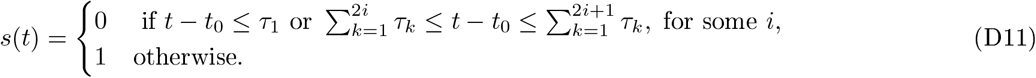

Here *t*_0_ is fixed and the variables 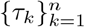 are assumed to be non-negative and add up to *t*_*f*_ − *t*_0_; this policy can hence be parametrized by a vector ***µ*** *∈* ℝ^*n*^, with ***τ*** = (*t*_*f*_ − *t*_0_)softmax(***µ***)^5^. Intuitively, 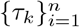 divide the interval [*t*_0_, *t*_*f*_] into *n* different parts. In each part, lockdown is alternatively declared (*s* = 1) or suspended (*s* = 0), see Figure 15.

**FIG. 15:**
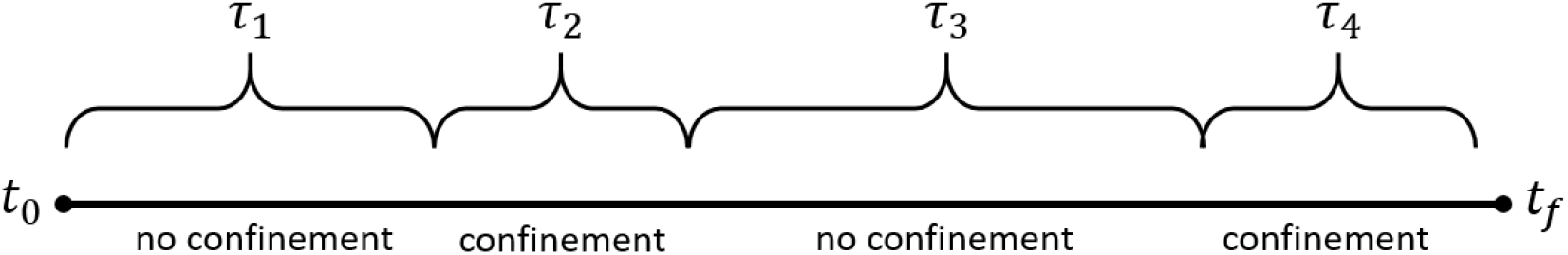
Parametrization of continuous lockdown times.

At time *t*, the disease’s basic reproduction number is given by (3), with *s*(*t*) defined as above. To find out 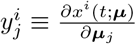, we invoke Eq. (C5). In computing the term

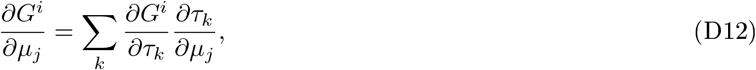

we have the problem that, due to (D11), *G* is not continuous or derivable. To work our way out, we approximate *s*(*t*) by a piece-wise continuous function with bounded derivative that transitions from 0 to 1 (or viceversa) linearly and in time *δ* ≪ 1, see Figure 16; later we will take the limit *δ →* 0.

**FIG. 16:**
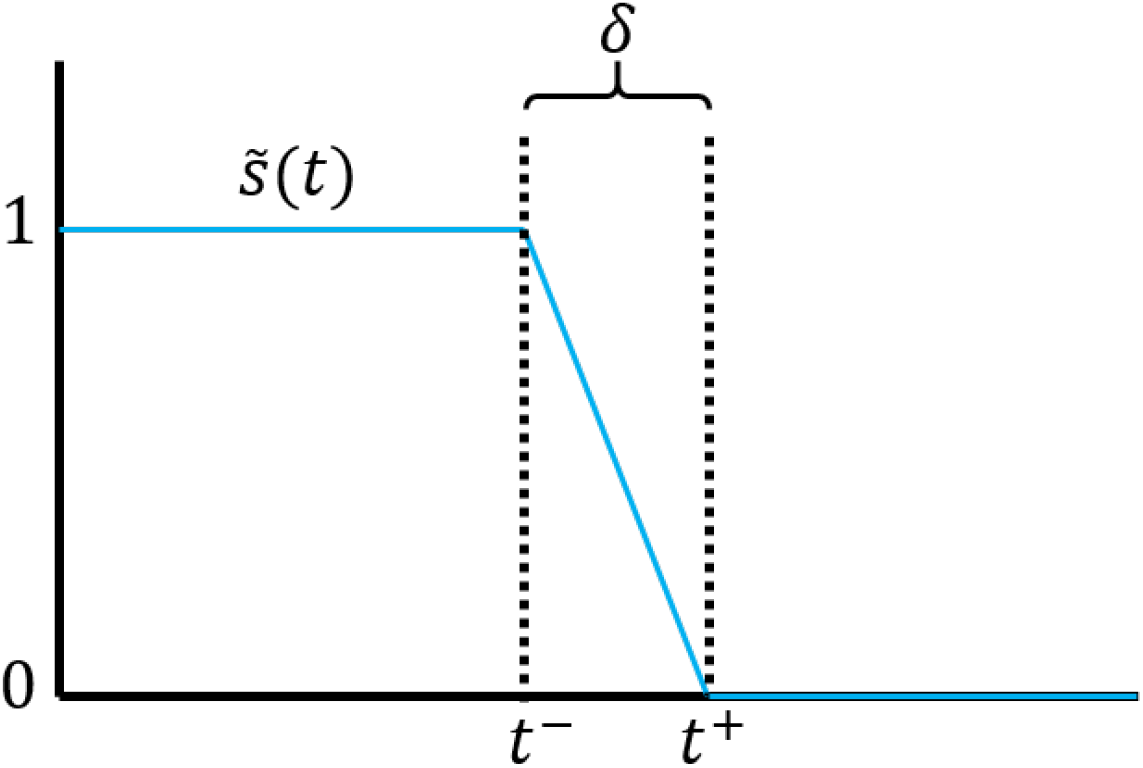
Modified continuous function 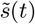.

The new function 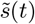 has zero derivative with respect to *µ*_*i*_, except for *t* satisfying

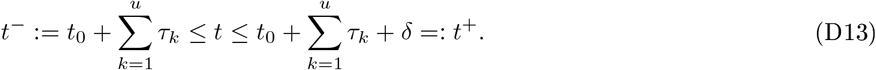

In that case, the derivative of *ŝ* with respect to *τ*_*j*_, with *j ≤ u*, will (approximately) be

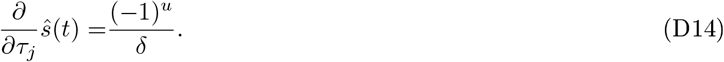

The derivative with respect to any of the variables *{τ*_*j*_ : *j > u}*−is zero.

The dominant term on the right-hand side of (C5) for *t ∈* [*t, t*^+^] is therefore

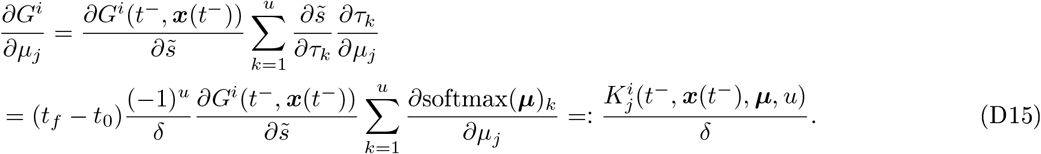

Since the evolution takes place for time *δ* = *t*^+^ − *t*^−^, we have that 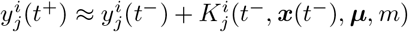. Taking the limit *δ →* 0, we have that the evolution of 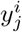 is determined by the following prescription:

1. 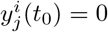.
2. Let 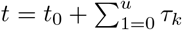, for some *u*. Then 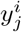 is updated by the rule

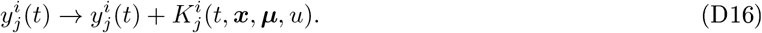
3. For all other values of 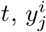 continuously evolves via the equation

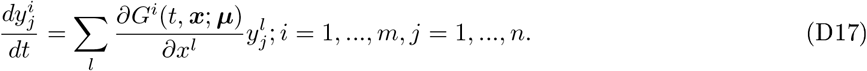

## APPENDIX E: CONTINUOUS-SPACE POPULATION MODELS

Even though the focus of this article is that of compartmental models of the form (C1), one can also apply the principles of gradient descent for policy optimizations on dynamical systems governed by partial differential equations. Consider, e.g., the scenario studied in [25], where the authors model the spread of rabies in raccoons across a realistic landscape Ω *⊂*ℝ ^2^through a system of reaction-diffusion equations of the form

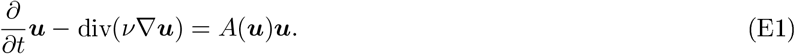

In this equation, the three entries of the vector field *u*(*t, X, Y*) *∈* ℝ^3^respectively denote the number of susceptible, exposed and infected individuals at time *t* in position *X, Y*. *ν, A* are 3 × 3 matrices that, in principle, might depend on some controllable parameters ***µ***. This equation is to be solved under the initial conditions *u*(0, *X, Y*) = *u*_0_(*X, Y*) and the homogeneous von Neumann boundary conditions

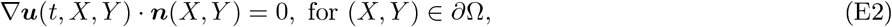

where ***n***(*X, Y*) *∈* ℝ^2^ denotes the vector normal to the contour *∂*Ω at location (*X, Y*). The authors of [25] solve this equation numerically via the Finite Element Method (FEM) [33].

Suppose that we wished to optimize the policy parameters ***µ*** *∈* ℝ^*n*^over some functional *A* depending on ***u***(*t, X, Y* ; ***µ***, *u*_0_) (instead of ***x***(*t*; ***µ, x***_0_)) via the gradient method. Then at some point we would need to compute the quantities 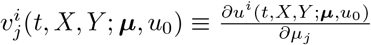. Call ***v***_***j***_ *∈* ℝ^3^the vector with components 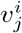 and differentiate both (E1) and (E2) with respect to *µ*_*i*_. This results in the equation

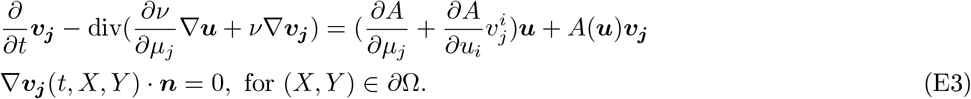

Since ***u***(0, *X, Y* ; ***µ, u***_**0**_) does not depend on ***µ***, this new diffusion equation must be solved for the initial conditions ***v***_***j***_(*X, Y*, 0) = 0. This can be achieved numerically in the same way that the authors of [25] solved Eq. (E1), that is, via the FEM.

## Footnotes

1 More precisely, in our model, the relevant compartment is *C*_*C*_(*t*), or the fraction of the population in critical care at instant *t*. This has to be compared with *B*_*c*_, the number of critical care beds per inhabitant. *𝒞* (t), B_c_ relate to *C*_*C*_ (*t*),*B*_c_ through the expressions *𝒞* (t) = *N*_*pop*_*C*_*C*_(t); *Bc* = *N*_*pop*_*B*_*c*_, where *N*_*pop*_ is the population size.

2 Rigorously speaking, stochastic gradient descent just minimizes the average value of the objective function *A*. By adding to *A* su_ciently strong penalties to the violation of each optimization constraint, we make sure that such constraints will hold with high probability when *x*_*0*_ and *ν* are sampled from the training distribution.

3 In the case of adaptive policies, some of such entries might also represent the components of the cell state θ [29], i.e., the internal variables used by the government to keep track of the evolution of the disease and guide future government interventions.

4 Θ(z) equals 1 for z ≥ 0, or 0, otherwise.

5 The function softmax(μ) of a vector μ is a vector *ν* of the same dimension with components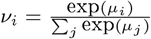

